# Quercetin enhances decidualization through AKT-ERK-p53 signaling and supports a role for senescence in endometriosis

**DOI:** 10.1101/2023.08.30.23294800

**Authors:** Julia Delenko, Xiangying Xue, Prodyot K Chatterjee, Nathaniel Hyman, Andrew J Shih, Robert P Adelson, Polona Safaric-Tepes, Peter K Gregersen, Christine N Metz

**Affiliations:** The Donald and Barbara Zucker School of Medicine, 500 Hofstra Blvd, Hempstead, NY 11549 USA; The Institute of Molecular Medicine, The Feinstein Institutes for Medical Research, Northwell Health, 350 Community Drive, Manhasset, NY 11030 USA

**Keywords:** decidualization, endometriosis, fertility, AKT signaling, p53 signaling, apoptosis, senescence

## Abstract

Quercetin, a flavonoid with senolytic activity, has attracted great interest as a therapy for fibrotic diseases such as pulmonary fibrosis, a disorder attributed to senescent pulmonary fibroblasts. Interestingly, quercetin has shown some benefit in pre-clinical models of endometriosis, an inflammatory condition characterized by senescent endometrial stromal cells and in severe cases, intraperitoneal fibrotic lesions and infertility. Quercetin exerts multiple biological activities but the signaling pathways underlying quercetin’s effects are not well-defined. In this report, we have analyzed the signaling pathways underlying quercetin’s action using menstrual effluent-derived endometrial stromal cells. We found that quercetin promotes decidualization, a well-defined differentiation process known to be defective in patients with endometriosis using cells obtained from endometriosis patients and unaffected controls. We show that quercetin substantially reduces the phosphorylation of multiple signaling molecules in the AKT and ERK1/2 pathways. In contrast, we observed striking phosphorylation of p53 and increased p53 protein expression. Furthermore, p53 inhibition blocks decidualization while p53 activation promotes decidualization. Finally, we provide evidence that quercetin increases apoptosis of endometrial stromal cells with a senescence phenotype. These data provide insight into mechanisms of action of quercetin in the setting of endometriosis and support studies to test senolytics for treating endometriosis.

## Introduction

Replicative and other forms of senescence have received wide attention as possible targets for the prevention of aging as well as potential treatments of chronic fibrotic disorders. For example, senolytic agents such as quercetin and dasatinib have shown promising results for pulmonary fibrosis in clinical trials (1, 2). In addition, these and other senolytic agents have been tested in numerous pre-clinical and pilot human studies and reported to be effective in a variety of other disorders characterized by chronic inflammation and fibrosis (3). As a common dietary flavonoid, the biological activity of quercetin has been widely studied, with evidence for anti-oxidant, anti-inflammatory, immune modulatory, anti-bacterial, anti-viral effects, as well as senolytic activities, among others. However, these effects vary among different cell types, and the exact signaling pathways have not been well-defined.

Reports in the literature support that quercetin may have therapeutic efficacy in animal models of endometriosis (4, 5). Additionally, in combination with other natural agents quercetin reduces pain in patients with endometriosis (6, 7). Endometriosis is a common, chronic inflammatory condition defined by endometrial-like lesions growing outside of the uterus, mainly in the pelvic cavity. It is characterized by chronic pelvic pain and often accompanied by dysmenorrhea, dyspareunia, dysuria, dyschezia, and infertility (8). The exact causes of endometriosis and associated infertility are not well understood. The delivery of menstrual tissues to the pelvic cavity is observed in nearly all menstruators (9) and, intriguingly, endometriosis is associated with altered endometrial tissue (shed as menstrual effluent, ME) (10–13). We have recently characterized ME-derived endometrial stromal cells (eSCs) in patients with endometriosis vs. unaffected controls (14, 15). In patients with endometriosis, these cells exhibit dramatic defects in decidualization (14–17), a common and essential differentiation event that is required for embryo implantation and reproductive success.

More recent single cell RNA-sequencing (scRNA-Seq) analysis of ME-eSCs by our group (18) supports altered differentiation of eSCs in the setting of endometriosis, as well as their deviation toward a more pro-inflammatory, pro-fibrotic, and senescent phenotype that may promote endometriosis lesion formation and/or disease progression. Based on these findings and the lack of effective pharmacological therapies for treating endometriosis, we assessed the effects of quercetin on this differentiation process, with the hypothesis that quercetin would improve decidualization and prevent differentiation of eSCs to cells with a pro-inflammatory and possibly senescent phenotype. After demonstrating that quercetin significantly enhanced decidualization responses using eSCs obtained from both control subjects as well as patients with endometriosis, we defined the signaling pathways involved in mediating quercetin’s effect and explored the potential role of senescence, which was previously shown to inhibit decidualization (19).

## Results

### Quercetin significantly reduces the proliferation of endometrial stromal cells

As described for many cell types, including uterine biopsy-derived eSCs (5), quercetin dose-dependently (12-50μM) reduced the proliferation of ME-derived eSCs obtained from healthy controls (Figure 1A).

**Figure 1.**
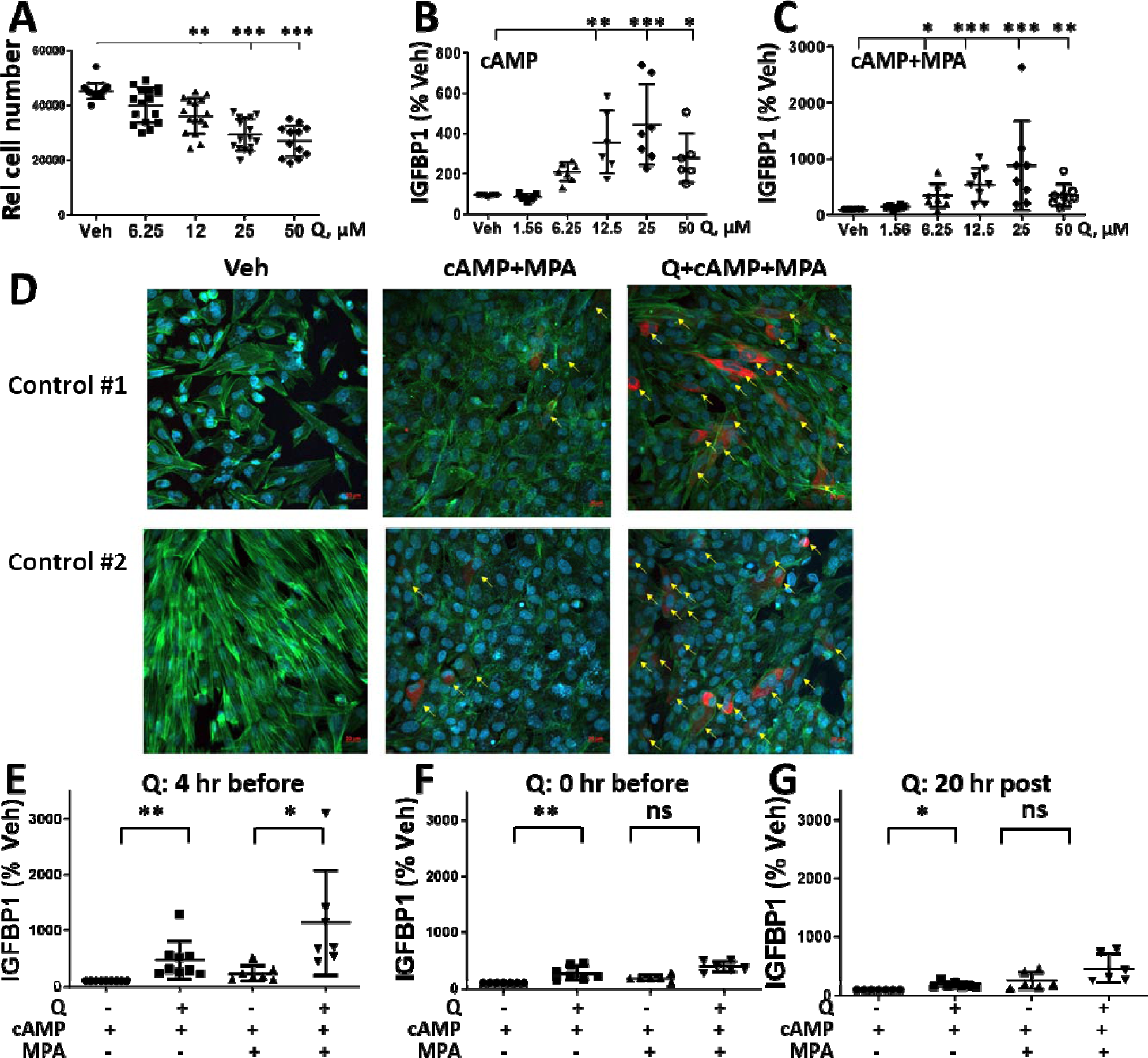
Quercetin inhibits eSC proliferation and enhances decidualization. **(A)** Control endometrial stromal cells (eSCs) were treated with vehicle or quercetin (0-50µM) for 72 hr and relative cell number (Rel cell number) was determined. **(B-C)** To determine the optimal dose of quercetin, control eSCs were treated with vehicle (Veh) or quercetin (0-50_μ_M) for 4 hr prior to the addition of cAMP alone (B) or cAMP+MPA (C). After 48 hours, decidualization was analyzed by measuring IGFBP1 levels by ELISA. Data are shown as IGFBP1 (percent Veh) where 100% = Veh-cAMP alone (IGFBP1) (B) or Veh-cAMP+MPA (C). **(D)** eSCs from two control subjects were treated with vehicle or quercetin (25µM) for 4 hr followed by vehicle or cAMP+MPA prior to staining and confocal imaging; images show IGFBP1 (red), phalloidin (green), and DAPI (blue) staining (at 20X magnification). The scale bars (20 µm) are indicated. IGFBP1+ cells are indicated by yellow arrows. Images showing all channels are in Supplementary Figure S1. **(E-G)** The effects of quercetin (25_μ_M) on decidualization when added 4 hr prior to cAMP±MPA (E) versus 0 hr before (at the time of) cAMP±MPA (F) or 20 hr post-cAMP±MPA stimulation (G). IGFBP1 levels were measured by ELISA 48 hr post cAMP±MPA stimulation. Data are shown as IGFBP1 (percent Veh) where 100% = Veh-cAMP alone (IGFBP1). Each point represents a datum from one individual’s eSCs, with mean (horizontal line) and SD (vertical lines) shown for each group. *p<0.05 vs. Veh, **p<0.01 vs. Veh; ***p<0.001 vs. Veh.

### Quercetin significantly enhances decidualization of cultured eSCs

The differentiation of eSCs into decidual cells is induced by progesterone (or its stable synthetic analogue, medroxyprogesterone acetate (MPA) and the cAMP/protein kinase A signaling pathway, which can be replicated in vitro using cell-permeable cAMP alone or cAMP+MPA (20). Testing a range of quercetin doses with eSCs obtained from healthy controls revealed that quercetin dose-dependently enhanced both cAMP- and cAMP+MPA-induced decidualization, as quantified by IGFBP1 production, peaking at 25μM quercetin (Figure 1B-C, respectively). IGFBP1 expression by control eSCs following cAMP+MPA-induced decidualization was confirmed by confocal microscopy (Figure 1D). IGFBP1 was absent without deciduogenic stimulation, and as previously described (21) not all cells express IGFBP1 following stimulation. Furthermore, quercetin increased cAMP+MPA-induced IGFBP1 expression by eSCs as determined by confocal microscopy (Figure 1D).

Using the optimal dose of quercetin (25µM), the most dramatic and consistent decidualization enhancing effects were observed when added 4 hr prior to cAMP or cAMP+MPA treatment (Figure 1E), rather than at the same time (Figure 1F) or following cAMP+MPA treatment (Figure 1G).

### Endometriosis-eSCs exhibit impaired decidualization when compared to control-eSCs

As previously reported (15), eSCs from endometriosis cases exhibit reduced decidualization response following cAMP alone (Figure 2A) and cAMP+MPA (Figure 2B) when compared to eSCs obtained from healthy controls, regardless of the decidualization stimulus (p<0.005). Note that eSCs from both cases and controls exhibit a range of decidualization responses (Figure 2).

**Figure 2.**
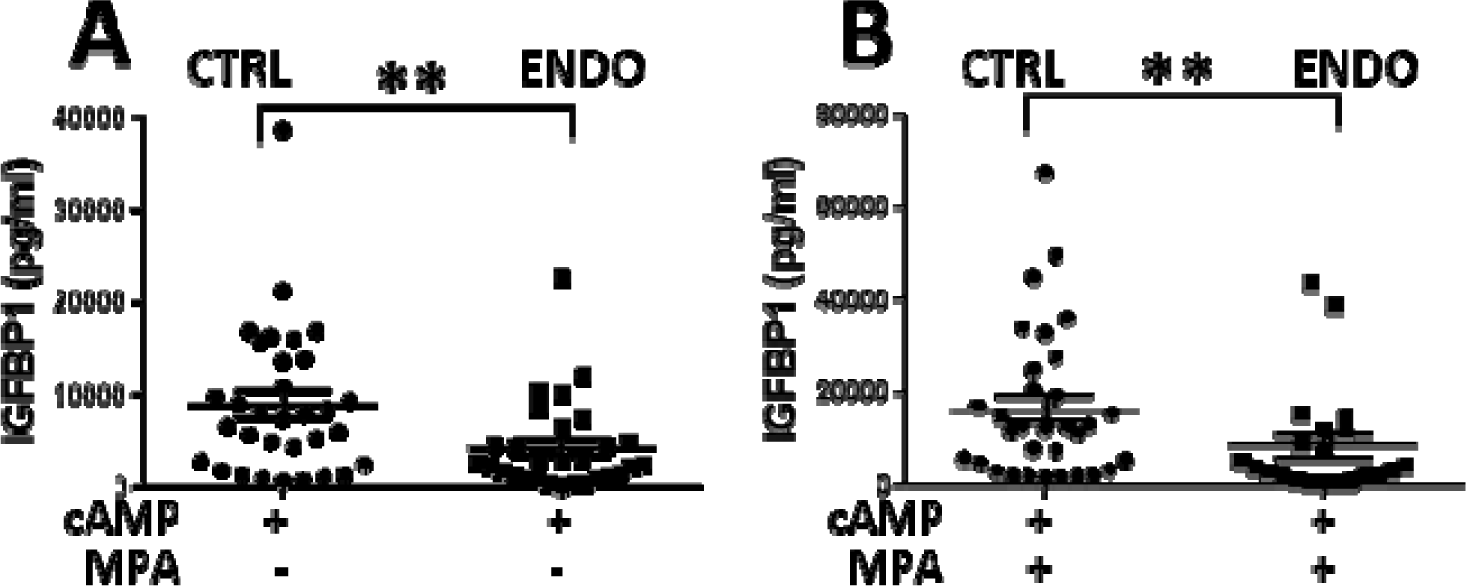
Endometriosis endometrial stromal cells (eSCs) exhibit impaired decidualization. **(A-B)** Comparison of decidualization responses of eSCs from controls (CTRL) vs. endometriosis (ENDO) cases induced by cAMP alone (A) or cAMP+MPA (B), as determined by IGFBP1 levels measured 48 hr post cAMP±MPA. Each point represents a datum from one individual’s eSCs, with mean (horizontal line) and SD (vertical lines) shown for each group. **p<0.001 CTRL vs. ENDO (Student’s t test).

### Quercetin promotes decidualization of eSCs from healthy controls and endometriosis cases

Using ME-derived eSCs isolated from healthy controls and endometriosis cases, quercetin treatment significantly enhanced both cAMP-induced (Figure 3A-B, p<0.0001) and cAMP+MPA-induced decidualization (as determined by IGFBP1 levels) (Figure 3C-D, p<0.0001) regardless of whether the donor was affected by endometriosis or not. Also, quercetin treatment of a subset of control-eSCs and endometriosis-eSCs significantly increased cAMP-induced and cAMP+MPA-induced production of PRL, another biomarker of decidualization (Figure 3E-H). Similar results were obtained for *IGFBP1* and *PRL* mRNA expression when analyzed by qPCR (Supplementary Figure S2).

**Figure 3.**
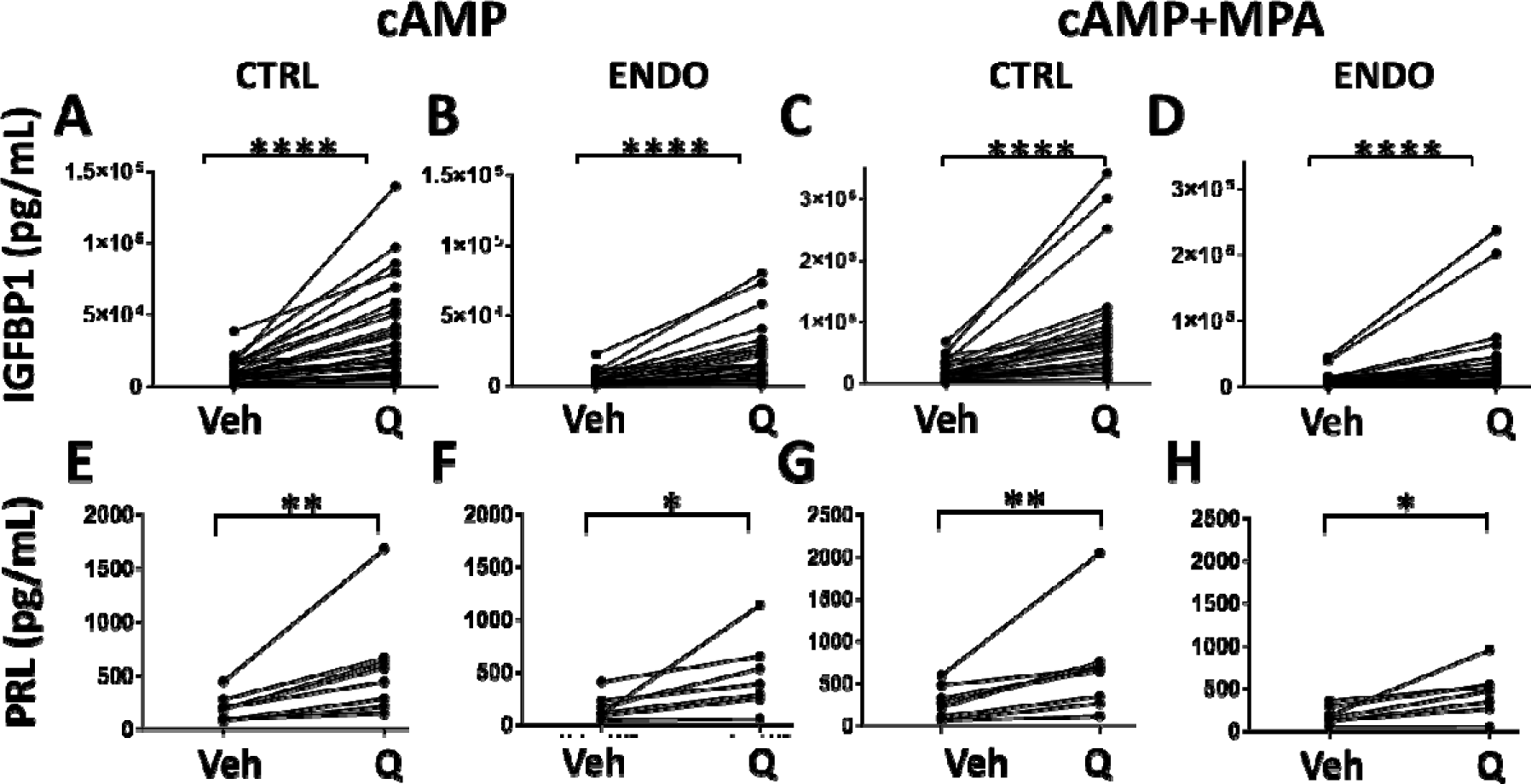
Quercetin enhances decidualization by control-eSCs and endometriosis-eSCs, as determined by IGFBP1 and PRL protein production. **(A-H)** Endometrial stromal cells (eSCs) from controls (A, C, E, and G, CTRL) and endometriosis patients (B, D, F, and H, ENDO) were treated with vehicle (Veh) or quercetin (Q, 25_μ_M) for 4 hr prior to the addition of cAMP alone (A-B, E-F) or cAMP+MPA (C-D, G-H). After 48 hr, decidualization was analyzed by measuring IGFBP1 (A-D) or PRL (E-H) levels by ELISA. Data points connected by a line represent paired mean data points from one individual’s eSCs (±Q). *p<0.05 Veh vs. Q-treated; **p<0.01 Veh vs. Q-treated; ****p<0.0001 Veh vs. Q-treated.

It has been proposed that prostaglandin E2 (PGE2) and progesterone are the minimal ancestral deciduogenic signals that induce decidualization in eutherian mammals (22). As shown for cAMP- and cAMP+MPA-induced decidualization (Figure 3), quercetin significantly elevates both PGE2- and PGE2+MPA-induced decidualization by control-eSCs (Supplementary Figure S3).

### Quercetin does not generate intracellular cAMP production by eSCs

Because cAMP drives decidualization and quercetin enhances intracellular cAMP ([cAMP]_i_) concentrations by the N1E-115 neuroblastoma cell line (23), we assessed the production of [cAMP]_i_ by quercetin-treated eSCs. This approach was supported by scRNA-Seq analyses revealing multiple *ADCY* transcripts (that encode adenylyl cyclases required to synthesize 3’,5’-cAMP) expressed by eSCs in fresh ME, including low expression of *ADCY1, 3, 6, 7,* and *9* transcripts (Supplementary Figure S4). Forskolin induced [cAMP]_i_ by control-eSCs (Figure 4A). However, quercetin treatment did not induce [cAMP]_i_ by eSCs (Figure 4A-B).

**Figure 4.**
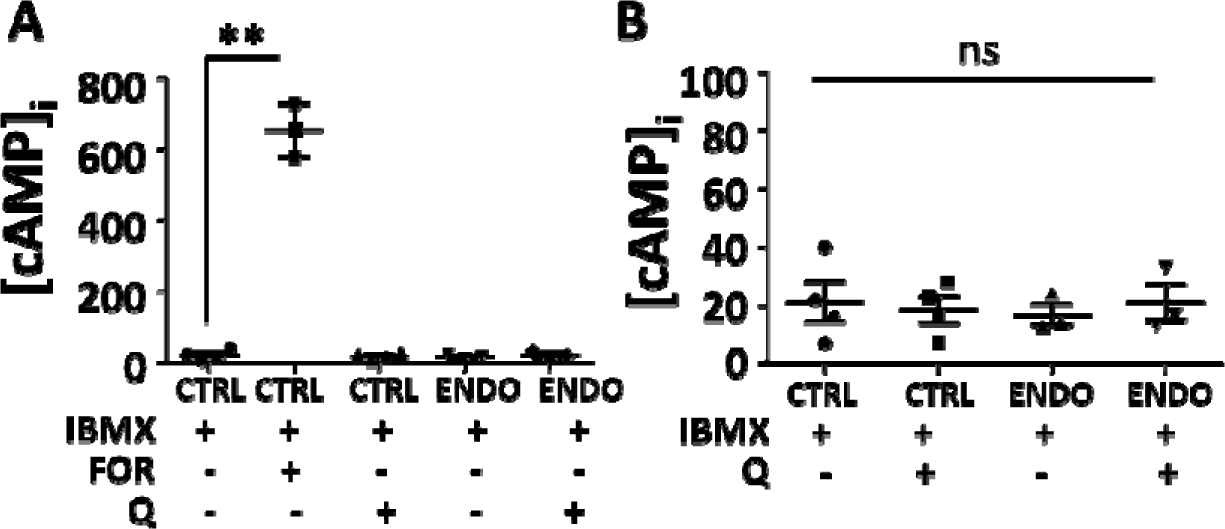
Quercetin does not increase [cAMP]_i_ concentrations in eSCs. (**A-B**) Endometrial stromal cells (eSCs) from controls (CTRL) or endometriosis cases (ENDO) were treated with IBMX (0.1mM), followed by addition of either vehicle, forskolin (FOR, 25µM) or quercetin (Q, 25µM). Lysates were analyzed for [cAMP]_i_ concentrations (pMol/mL) (A-B). A subset of data are shown on a different scale without forskolin treatment (B). Each point represents a datum from one individual’s eSCs, with mean (horizontal line) and SD (vertical lines) shown for each group. **p<0.001 vs. IBMX; ns=not significant.

### Quercetin does not exert anti-oxidant effects in eSCs

Measuring oxidative stress using the DCFDA assay, H_2_O_2_-treatment significantly induced oxidative stress in both control and endometriosis-eSCs (Figure 5). While the well-known antioxidant, *N*-acetylcysteine (NAC), significantly reduced H_2_O_2_-induced oxidative stress by eSCs (Figure 5A-D), quercetin did not reduce oxidative stress by eSCs when added <u>after</u> (Figure 5A-B) or <u>before</u> H_2_O_2_ stimulation (Figure 5C-D).

**Figure 5.**
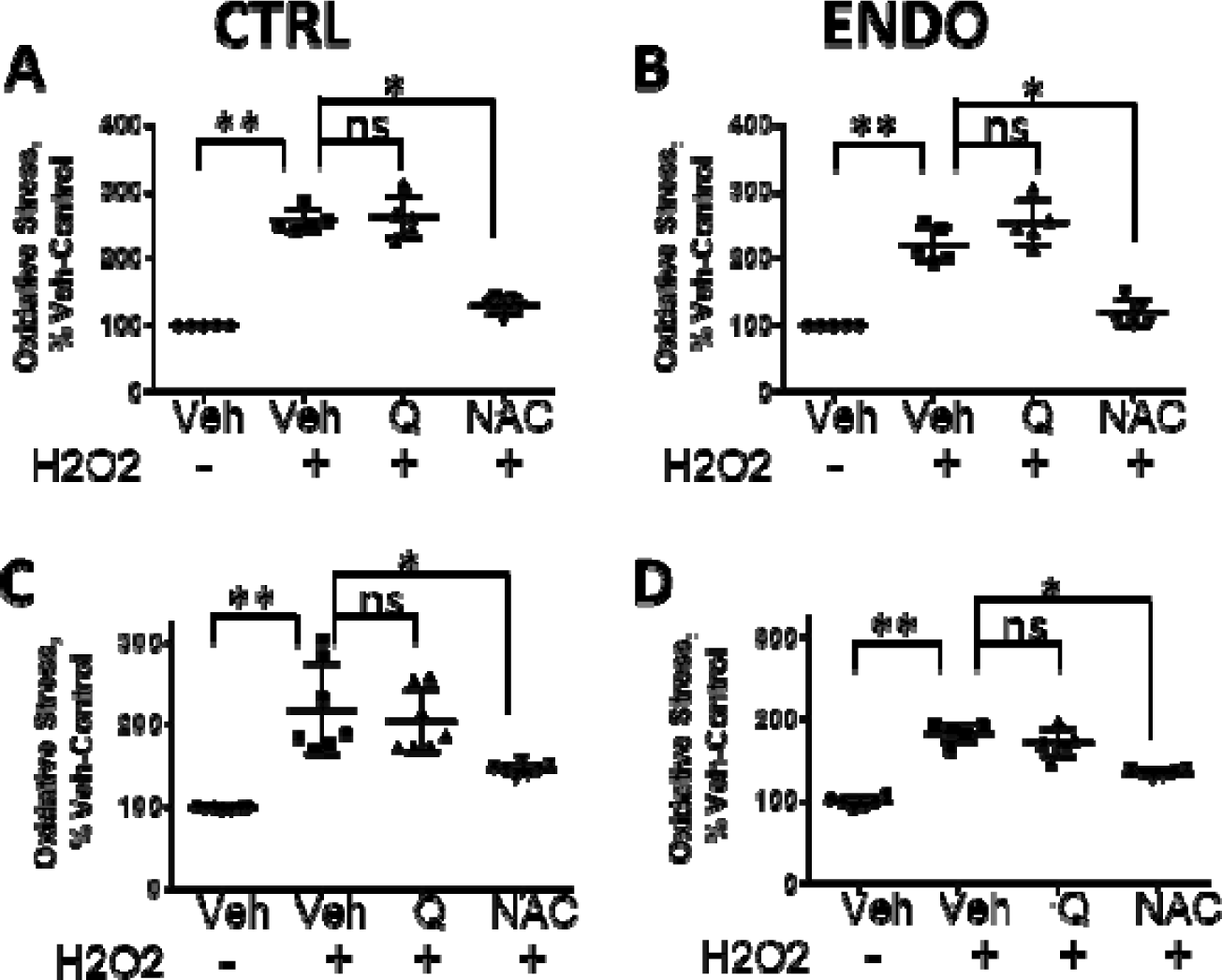
Quercetin does not reduce oxidative stress in eSCs. **(A-D)** Endometrial stromal cells (eSCs) from controls (CTRL, A and C) or endometriosis cases (ENDO, B and D) were treated with either vehicle (Veh), quercetin (Q, 25_μ_M), or *N*-acetylcysteine (NAC, 10mM) after H_2_O_2_ stimulation (A and B) or before H_2_O_2_ stimulation (C and D), and oxidative stress was measured using the DCFDA assay. Data are shown as percent control oxidative stress where 100% = Veh-Veh-treated eSCs (oxidative stress). Each point represents a datum from one individual’s eSCs, with mean (horizontal line) and SD (vertical lines) shown for each group. *p<0.05; **p<0.01; and ns = not significant.

### Quercetin treatment of eSCs alters numerous signaling pathways

Phospho-kinase arrays were used to investigate various signaling pathways altered by quercetin. Compared to vehicle treatment of control-eSCs, quercetin consistently reduced the expression of phospho-AKT (T308 and S473), phospho-ERK1/2 (multi), phospho-PRAS40 (T246), and phospho-WNK1 (T60), and significantly increased phospho-p53 (S46) expression, when assessed 4 hr after treatment (Figure 6A-B, Supplementary Figure S5).

**Figure 6.**
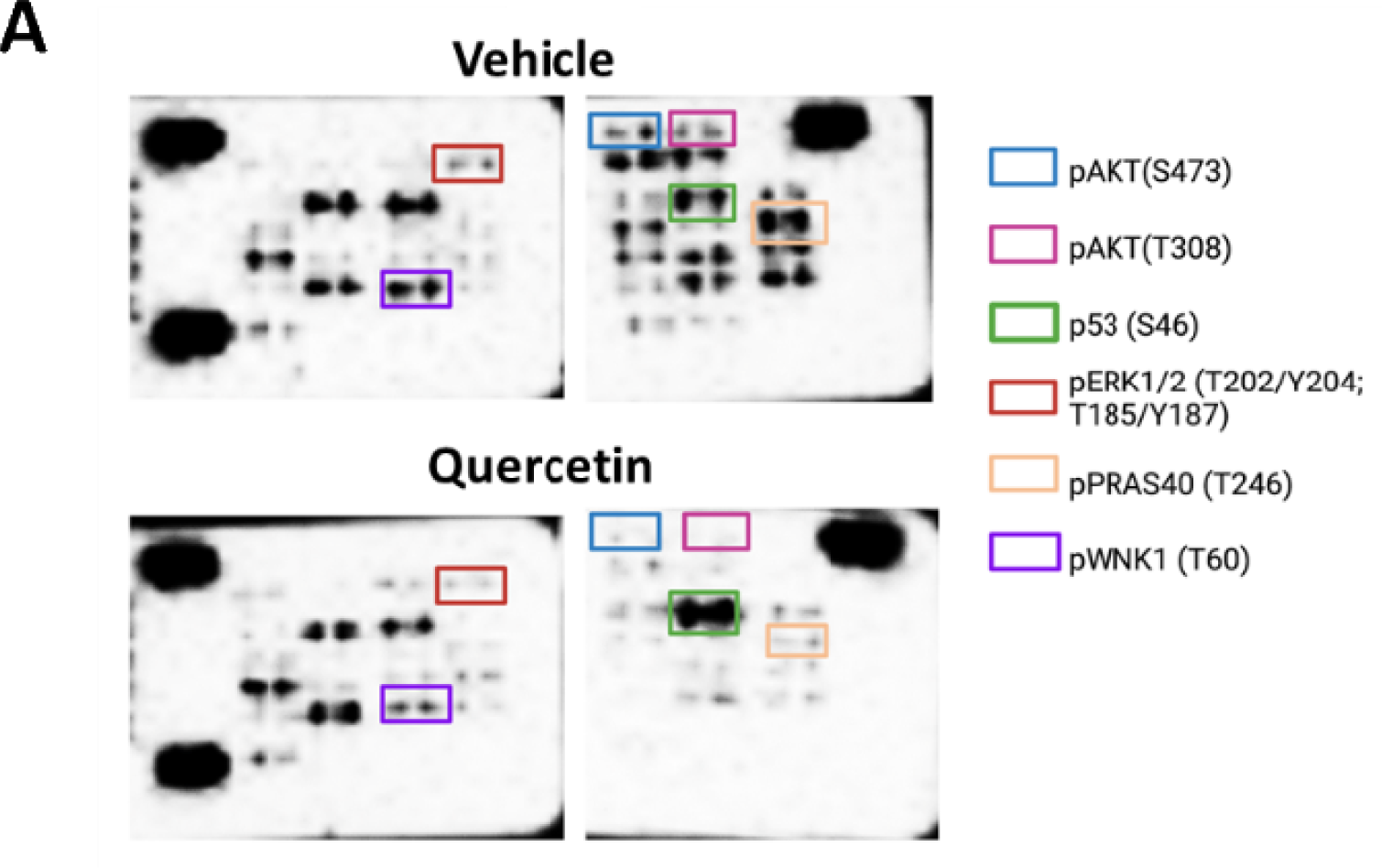
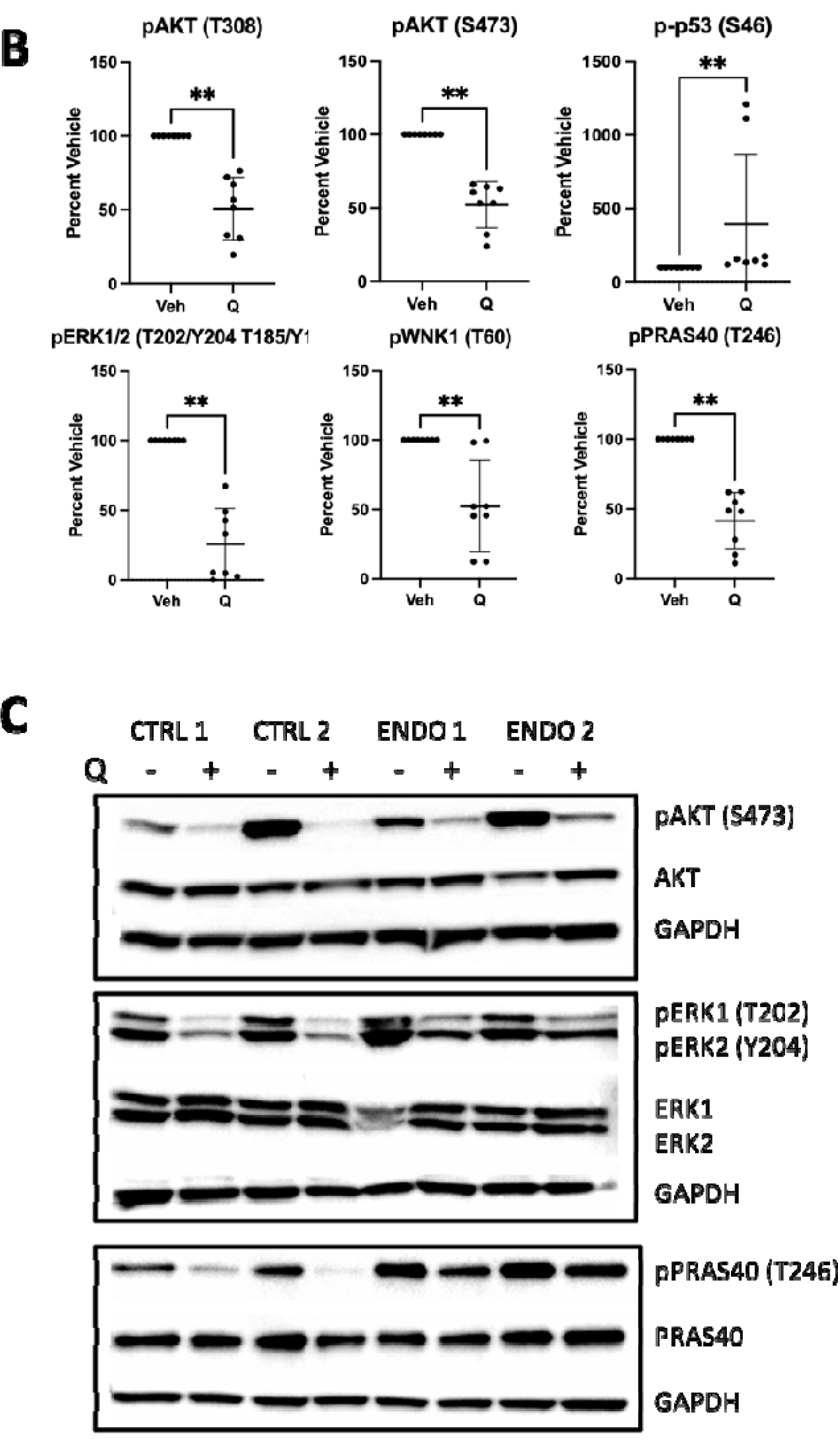
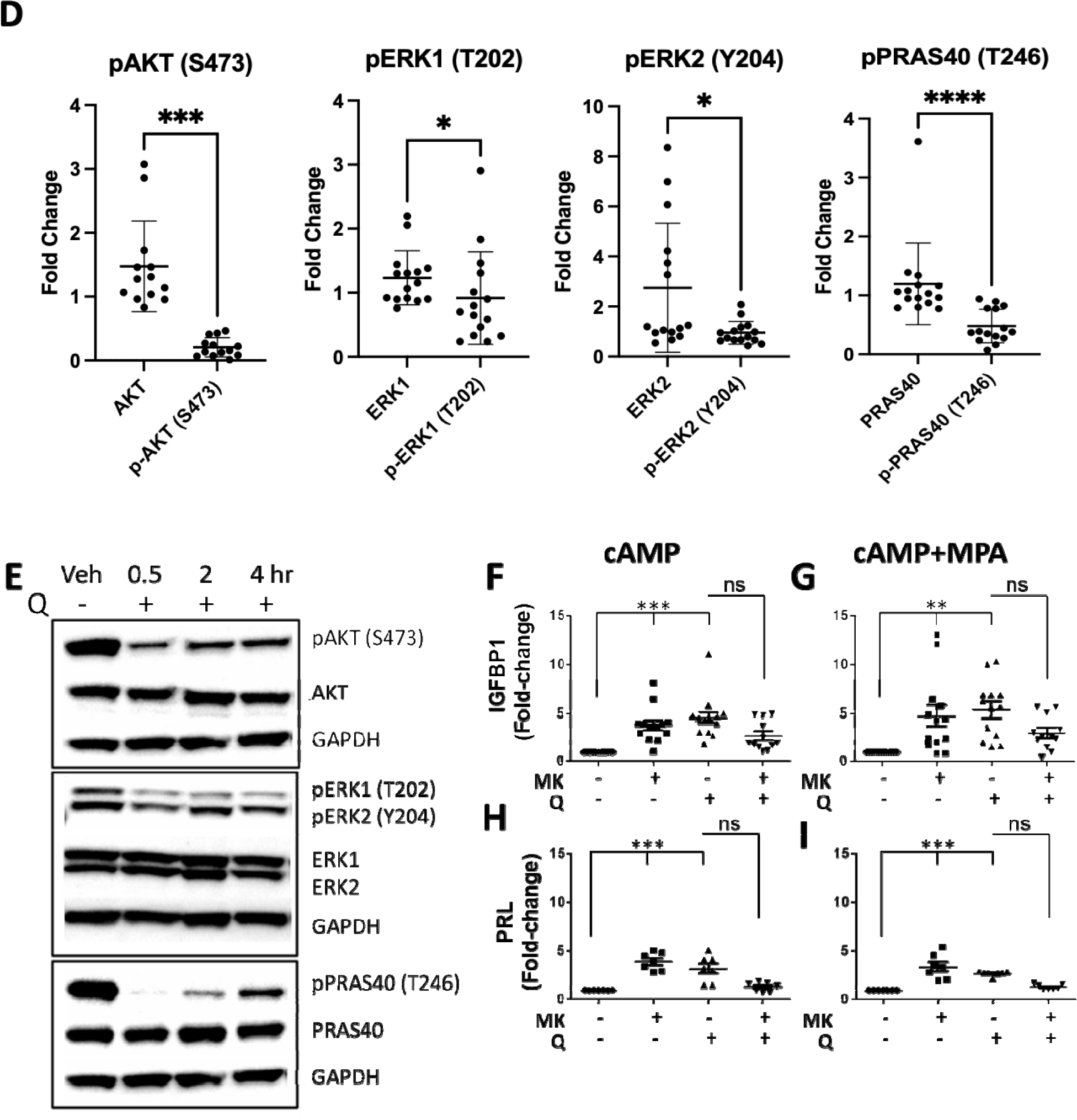
Quercetin inhibits AKT and ERK1/2 phosphorylation and signaling and promotes p53 (Ser46) phosphorylation. **(A-B)** Control endometrial stromal cells (eSCs) were treated with vehicle (Veh) or quercetin (Q, 25_μ_M) for 4 hr before lysing cells and analyzing cell lysates using a phospho-kinase array panel. Representative arrays and quantification of array analytes are shown in (A) and (B), respectively. For quantification, data for differentially expressed analytes are presented as analyte density normalized to the reference spot comparing Veh-treated vs. Q-treated. Each point represents the specific density of each analyte spot as a percentage of Veh-treated, with mean (horizontal line) and SD (vertical lines) shown for control- and endometriosis-eSCs (the remaining arrays are shown in Supplementary Figure S5). **p<0.01. **(C-D)** Control (CTRL) and endometriosis (ENDO) eSCs were treated with Veh or Q (25µM) for 4 hr before western blot analysis of cell lysates for p-AKT, total AKT, p-ERK1/2, total ERK1/2, p-PRAS40, and total PRAS40. Representative blots and quantification of specific analytes are shown in (C) and (D), respectively. Band densities, shown as fold-change between Veh-vs. Q-treated eSCs in (D), are normalized to the specific non-phosphorylated protein. Each point represents a datum from one individual’s eSCs, with mean (horizontal line) and SD (vertical lines) shown for each group *p<0.05; ***p<0.0001; ****p<0.00001; ns=non-significant. (**E)** Control-eSCs were treated with Veh or Q (25µM) for 0.5, 2, and 4 hr before western blot analysis of cell lysates for phospho-AKT, total AKT, phospho-ERK1/2, total ERK1, phospho-PRAS40, and total PRAS40. **(F-I)** Control-eSCs were treated with vehicle (Veh), AKT inhibitor MK-2206 (MK, 1μM), quercetin (Q, 25μM) or Q+MK for 4 hr prior to cAMP (F and H) or cAMP+MPA (G and I) stimulation; decidualization was assessed by measuring IGFBP1 levels (F and G) or PRL (H and I) by ELISA. Data are presented as fold-change in IGFBP1 or PRL over Veh-treated (Veh-treated=1). Each symbol represents a datum from one individual’s eSCs, with mean (horizontal line) and SD (vertical lines) for each group. **p<0.01; ***p<0.0001; ns=non-significant.

We confirmed that quercetin decreased the expression of phospho-AKT (S473), phospho-ERK1/2 (multi), and phospho-PRAS40 (T246) when analyzed by western blotting 4 hr later using control-eSCs and endometriosis-eSCs without changing total unphosphorylated target protein expression (Figure 6C-D). Time course analyses reveal that quercetin reduced phospho-AKT (S473), phospho-ERK1/2 (multi), and phospho-PRAS40 (T246) by control-eSCs within 30 min and this downregulation persisted for 4 hr (Fig. 6E). Accordingly, MK-2206, the highly selective inhibitor of AKT1/2/3, significantly enhanced both cAMP-induced IGFBP1 and PRL production (Figure 6F and 6H, respectively) and cAMP+MPA-induced IGFBP1 and PRL production (Figure 6G and 6I, respectively). The effect of MK-2206 on decidualization was similar to that achieved by quercetin and when MK-2206 was combined with quercetin, no additive effects were observed (Figure 6F-I), supporting a similar or overlapping pathway.

### Quercetin increases p53 protein expression

Although quercetin-induced phospho-p53 (Ser45) expression was observed using the phospho-kinase array (Figure 6A-B), it was not detected by standard western blotting methods likely due to the lower detection sensitivity for western blotting when compared to the array’s capture method. Regardless, quercetin increased total p53 protein levels by control-eSCs and endometriosis-eSCs after 4 hr (Figure 7A-B). Although higher p53 protein levels did not persist for 2 days (Figure 7C-D), the increase in total p53 protein was detected as early as 2 hr post-quercetin addition (Figure 7E). Consistent with these observations, pifithrin (PFT, 40µM), an inhibitor of p53, significantly reduced cAMP+MPA-decidualization when compared to vehicle (Figure 7F) and caused a similar reduction even with quercetin-enhanced decidualization (Figure 7G). By contrast, nutlin-3a, an inhibitor of MDM2 and promoter of p53, improved decidualization when added post-cAMP+MPA (Figure 7H).

**Figure 7.**
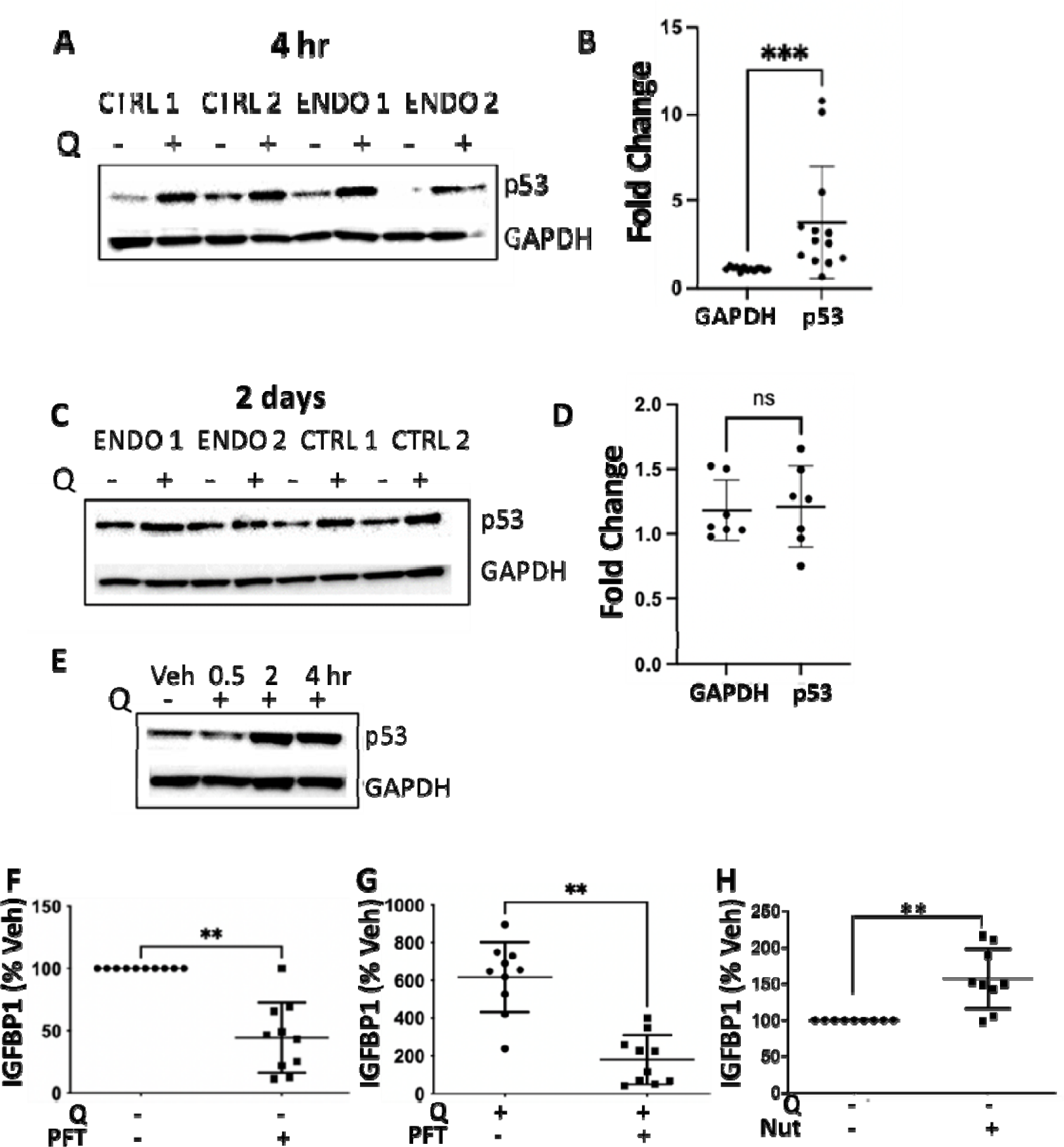
Quercetin induces p53, which contributes to decidualization. **(A-D)** Control (CTRL)- and endometriosis (ENDO)-endometrial stromal cells (eSCs) were treated with vehicle or quercetin (Q, 25 µM) for 4 hr (A, B) or 2 days (C, D) before western blot analysis of cell lysates for p53 and GAPDH. Representative blots are shown in (A) and (C). Band densities for control-eSCs were normalized to GAPDH; data are presented as fold-change after 4 hr (B) or 2 days (D) following quercetin treatment. **(E)** Control-eSCs were treated with Veh or Q (25 µM) for 0.5, 2, and 4 hr before western blot analysis of cell lysates for p53 and GAPDH. (**F-G)** Control-eSCs were treated with vehicle or Pifithrin _α_ (PFT, 40 _μ_M) for 4 hr prior to cAMP+MPA stimulation (F) or PFT (40 _μ_M) and Q (25_μ_M) for 4 hr prior to cAMP+MPA stimulation (G). Decidualization was assessed by measuring IGFBP1 levels by ELISA 48 hr later. **(H)** Control-eSCs were treated with Veh or nutlin-3a (Nut, 100nM) 24 hr after cAMP+MPA stimulation and decidualization was assessed by measuring IGFBP1 production 48 hr post-cAMP+MPA. For F-H, data are presented as IGFBP1 percent vehicle (Veh-treated cAMP=100%). Each point represents a datum from one individual’s eSCs, with mean (horizontal line) and SD (vertical lines) shown for each group. **p<0.01 vs. Veh-treatment.

### eSCs show evidence of a senescence-like phenotype, which is reduced by quercetin treatment

Our recent studies revealed numerous senescence-related gene markers that were consistently upregulated in eSCs of fresh ME tissues obtained from endometriosis patients vs. controls (18). Under basal conditions, early passage eSCs (p2-p3) express SASP biomarkers IL-6 and MMP3 (Figure 8A), and this SASP phenotype is reduced by quercetin treatment (Figure 8A-C). Also under basal conditions, early passage control-eSCs (p2-p3) express senescence-related markers p21 and p16 (which increase with senescence), and lamin B1 (which decreases with senescence) (Figure 8D). As expected, low dose H_2_O_2_, which induces eSC senescence (24), further induced p16, p21, and MMP3 expression and reduced lamin B1 expression over time (Figure 8D). Also, H_2_O_2_ treatment dramatically reduced decidualization (Figure 8E-F) and IGFBP1 levels were significantly restored by quercetin treatment prior to decidualization – albeit in a blunted manner when compared to its effect in the absence of H_2_O_2_ (Figure 8F).

**Figure 8.**
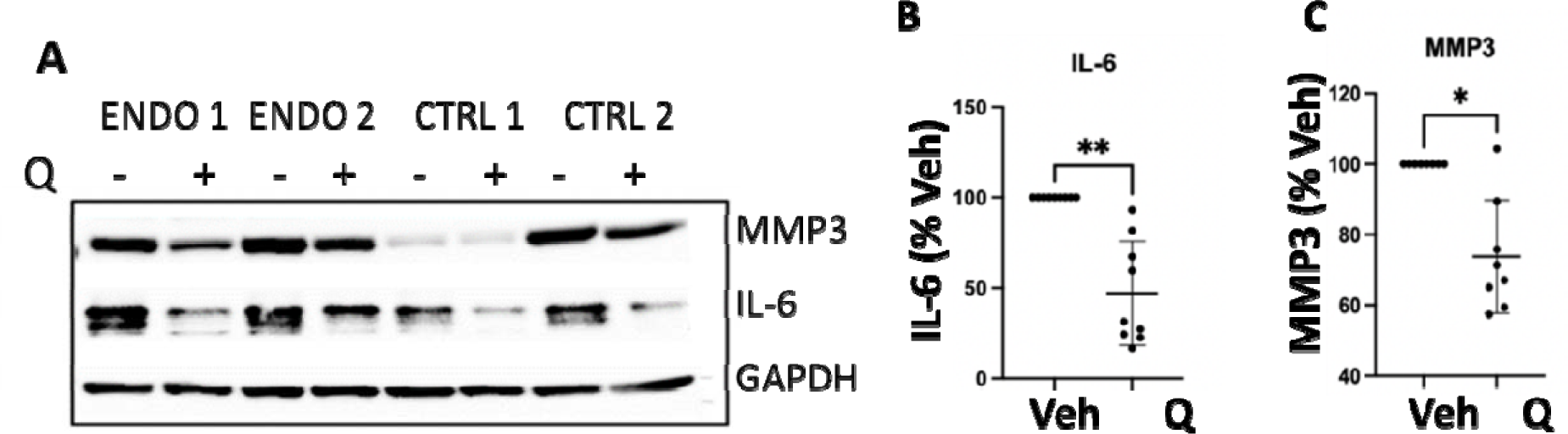
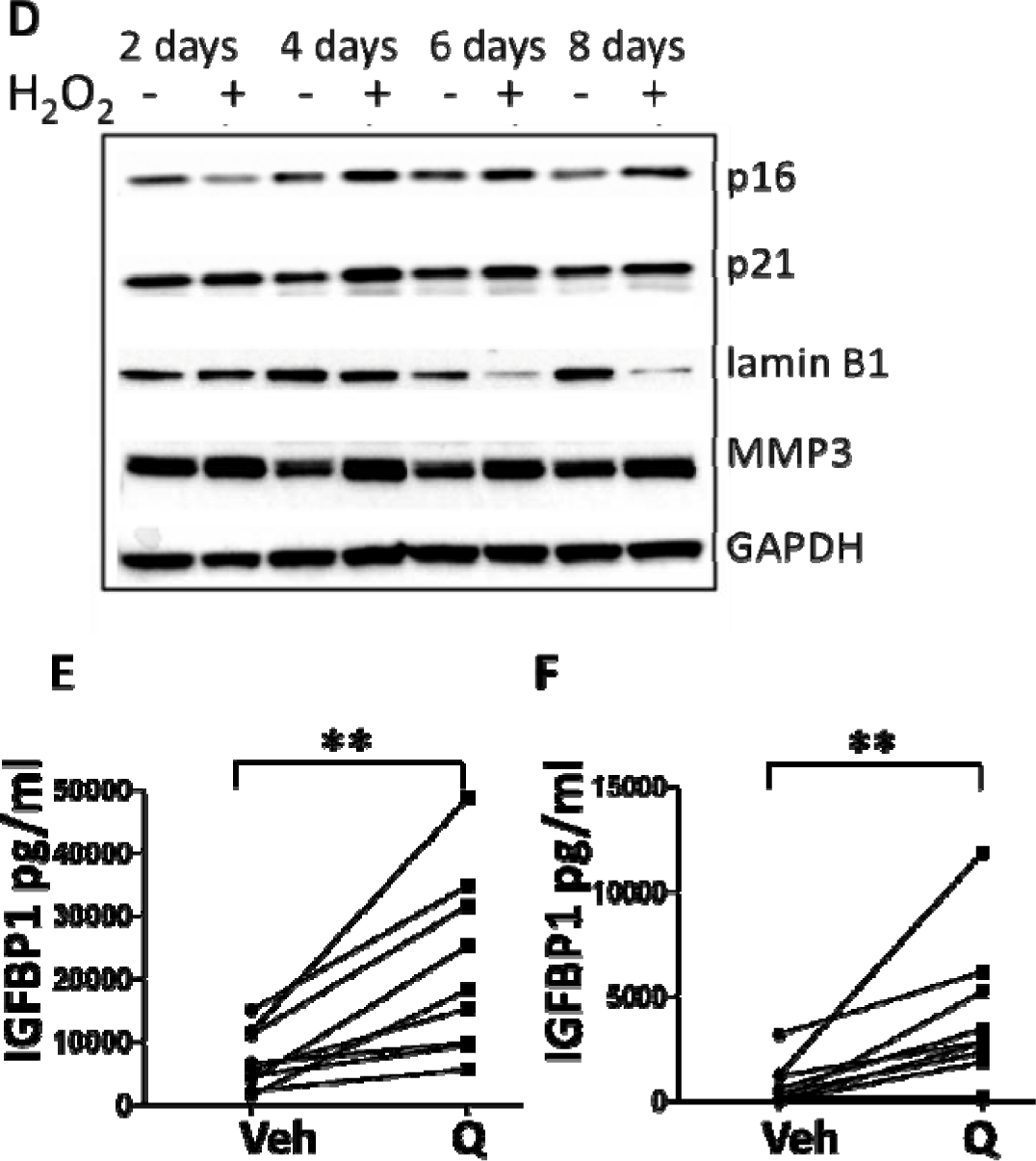
eSCs show evidence of senescence phenotype, which is reversed by quercetin. **(A)** Endometriosis (ENDO)-endometrial stromal cells (eSCs) and control (CTRL)-eSCs were treated with vehicle or quercetin (Q, 25 µM) for 2 days before western blot analysis of lysates for SASPs, IL-6 and MMP3, or GAPDH. A representative blot is shown. **(B-C)** Quantification of blots (with band densities normalized to GAPDH) from control-eSCs treated with vehicle (Veh) or quercetin (Q, 25 µM) for 2 days for IL-6 and MMP3 are shown in (B) and (C), respectively. Data are presented as IL-6 percent vehicle or MMP3 percent vehicle (where Veh-treated=100%). Each point represents a datum from one individual’s eSCs, with mean (horizontal line) and SD (vertical lines) shown for each group *p<0.05; ns=non-significant. **(D)** Control-eSCs were treated with vehicle or 250 µM H_2_O_2_ for 2 hr and then, eSCs were harvested at 2, 4, 6, and 8 days post-H_2_O_2_ exposure for western blot analysis for p21, p16, lamin B1, MMP3, and GAPDH. **(E-F)** Control-eSCs were treated with vehicle (E) or 250 µM H_2_O_2_ for 2 hr (F) and then treated with vehicle (Veh) or quercetin (Q, 25 µM) for 4 hr prior to cAMP+MPA-induced decidualization. After 48 hr, culture supernatants were assessed for IGFBP1 by ELISA. Data points connected by a line represent paired mean data points from one individual’s eSCs (±Q). *p<0.05 Veh vs. Q-treated; **p<0.01 Veh vs. Q-treated.

### Inhibition of apoptosis blocks decidualization; quercetin induces apoptosis in a subset of eSCs

Based on the role of p53 (S46) in apoptosis and the increase in p53 expression by quercetin, we tested the effect of Z-VAD-fmk, a pan-caspase inhibitor that prevents apoptosis, on decidualization. Pretreating control-eSCs with Z-VAD-fmk (40 µM) significantly reduced decidualization and blocked quercetin-enhanced decidualization when compared to vehicle-treated eSCs (Figure 9A-B). Accordingly, quercetin treatment significantly increased eSC apoptosis in a subset of eSCs when analyzed 24 and 48 hr later (Figure 9C-D). In addition, quercetin treatment decreased the number of large beta-galactosidase^+^ cells (Figure 9E-F), which were likely senescent.

**Figure 9.**
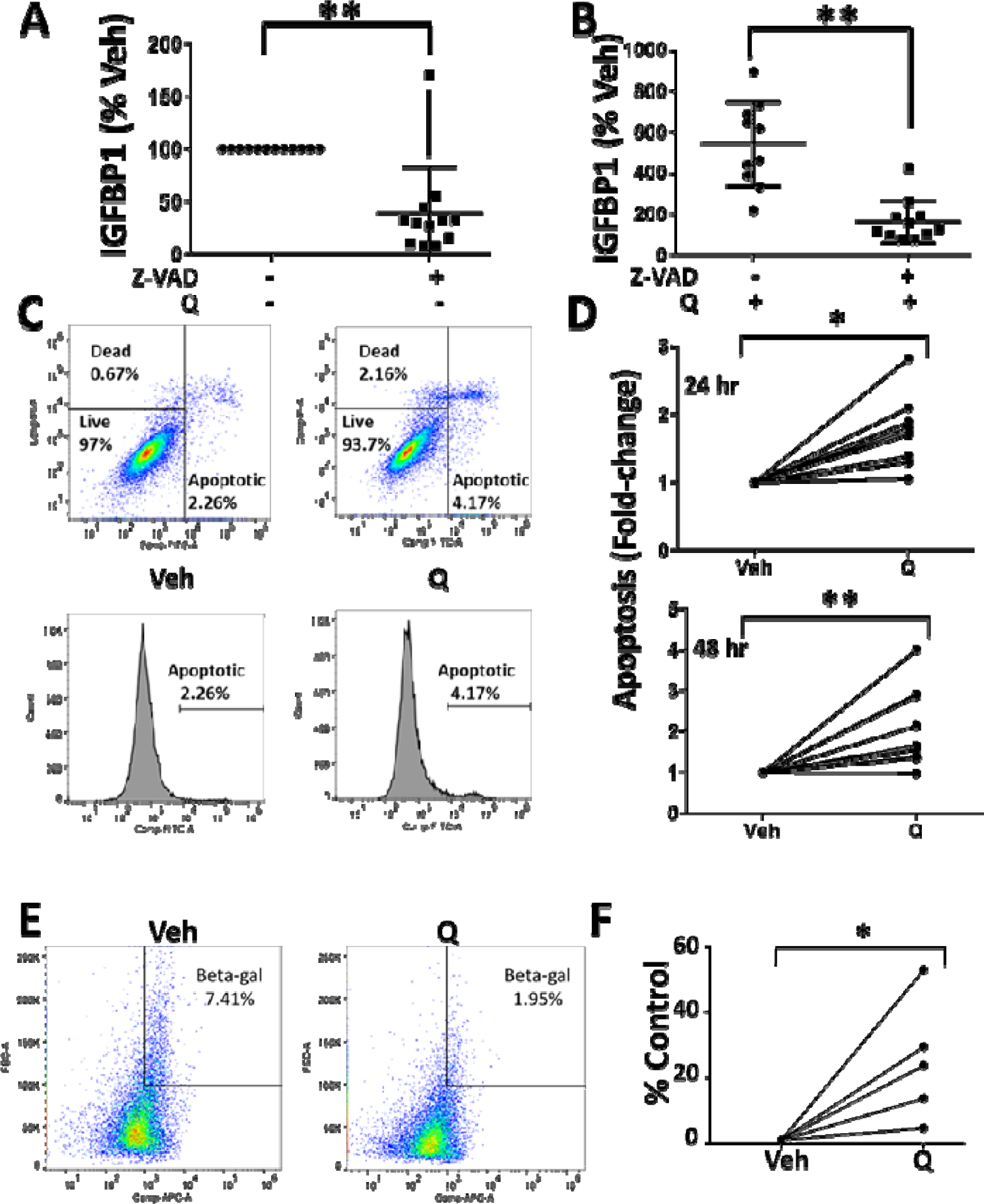
Inhibiting apoptosis blocks decidualization and quercetin induces apoptosis in a subset of eSCs. **(A-B)** Control-endometrial stromal cells (eSCs) (p3) were treated with vehicle (Veh) or Z-VAD-fmk (0 vs. 40 µM) in the absence of quercetin, i.e., vehicle (Veh) (A), or in the presence of quercetin (Q, 25 µM) (B) prior to inducing decidualization with cAMP+MPA. Decidualization was assessed 48hr post-cAMP+MPA treatment by ELISA for IGFBP1. Each point represents a datum from one individual’s eSCs, with mean (horizontal line) and SD (vertical lines) shown for each group. **p<0.01 comparing vehicle vs. Z-VAD-fmk. (**C-D**) Quercetin induces apoptosis in a subset of control-eSCs (passage 3-4). Representative gating (upper panels) and histograms (lower panels) of Annexin V staining (apoptosis) comparing vehicle-treated (Veh – left panels) vs. quercetin-treated (Q, 25 µM-right panels) eSCs 24 hr post-treatment (C). Quercetin induces apoptosis in various control-eSCs, shown by fold-change in apoptosis as determined by Annexin V staining 24 hr and 48 hr after vehicle (Veh) vs. quercetin (Q, 25 µM) treatment (D). Data points (fold-change in apoptosis ±Q) connected by a line represent paired mean data points from one individual’s eSCs (±Q). *p<0.05; **p<0.01 comparing vehicle vs. Q. (**E-F**) Larger senescence-associated β-galactosidase-positive eSCs are depleted following quercetin treatment. Representative gating of larger senescence-associated β-galactosidase (Beta-gal) positive eSCs comparing vehicle-treated (Veh – left panel) vs. quercetin-treated (Q, 25 µM-right panel) eSCs (passage 3-4) 48 hr post-treatment (E). Senescent cells were defined by high fluorescent-based β-galactosidase staining and large cell size, as measured by forward scatter (FSC-A). The percentage of senescent cells (of total viable cells per eSC sample) is indicated on each plot. The change in the number of larger senescence-associated β-galactosidase-positive control-eSCs 48 hr after treatment with vehicle (Veh) vs. quercetin (Q, 25 µM) is shown as percent control (control=vehicle-treated) (F). Data points connected by a line represent paired mean data points from one individual’s eSCs (±Q). *p<0.05 comparing vehicle vs. Q.

## Discussion

The data in this report provide mechanistic insight as to how quercetin enhances the decidualization response of eSCs. Decidualization refers to the differentiation of fibroblast-like eSCs into enlarged specialized decidual stromal cells that produce the growth factors needed to create a nutrient-rich uterine environment that is critical for implantation and successful pregnancy (20). In mice and most other eutherian mammals this process occurs post-implantation. By contrast, decidualization occurs spontaneously during each secretory phase of the menstrual cycle in the 4% of mammals that menstruate, including humans.

It is well-established that progesterone raises [cAMP]_i_ levels to trigger decidualization in vivo and this process can be recapitulated with cultured eSCs using cell-permeable cAMP ± MPA (20). Using this model system, numerous studies report decidualization defects with uterine biopsy-derived eSCs (16, 17, 25) and ME-derived eSCs (14, 15) in the setting of endometriosis. Here, we report that quercetin enhanced the decidualization response regardless of whether eSCs were obtained from endometriosis cases or unaffected healthy controls (Figure 3). However, quercetin did not increase [cAMP]_i_ levels (Figure 4), nor did it appear to mediate decidualization through anti-oxidant activity (Figure 5).

Previous studies have linked quercetin’s effect on decidualization to its anti-proliferative effect (4, 5, 26). However, not all agents that inhibit eSC proliferation enhance decidualization responses, suggesting that this effect is likely mediated through specific signaling pathways. We now show that quercetin rapidly reduces the phosphorylation of AKT (S473/T308), as well as ERK1/2 (multi), PRAS40 (T246), and WNK1 (T60). In contrast, quercetin enhances signaling in the p53 pathway, with increased phosphorylation of p53 (S46) and increased expression of total p53 protein.

These data are consistent with prior studies revealing AKT dephosphorylation during decidualization and increased phospho-AKT expression by stromal cells in the eutopic endometrium of endometriosis cases and endometriosis lesions (27–29). We propose that persistent phospho-AKT activity may explain decidualization defects observed in the setting of endometriosis that can be corrected with quercetin, since quercetin rapidly reduced AKT phosphorylation by eSCs (within 30 min, Figure 6E). Likewise, the AKT inhibitor, MK-2206, significantly increased decidualization (Figure 6F-G). AKT is a pro-survival factor with potent apoptosis-inhibiting activity whose function in regulating transcription factors and other proteins is mediated, in part, by its phosphorylation status. Thus, our findings are consistent with the ‘AKT inhibitory action’ of quercetin on eSCs (5) and other cells (30) and may explain the connection between inhibition of AKT signaling and increased decidualization and fertility (31)^;^(32).

A prior study reported that quercetin reduced *TP53* mRNA expression by primary eSCs while promoting decidualization (26). By contrast, we observed that quercetin increased p53 phosphorylation (Figure 6A-B) and total p53 protein levels (Figure 7A-B). We did not assess *TP53* mRNA expression and *TP53* mRNA expression may not correlate with p53 protein expression and activation. Our data support that the effects of quercetin on p53 expression and activation are related to the AKT and ERK1/2 pathways. AKT phosphorylates numerous proteins, including ‘with no lysine kinase-1’ or WNK1 (T60) (33) and ‘proline-rich AKT substrate of 40 kDa’ (PRAS40), which in turn enhances activation of PI3K/AKT signaling (34).

Accordingly, we found that quercetin reduced the phosphorylation of PRAS40 and WNK1 and by eSCs within 30 min (Figure 6E). Interestingly, PRAS40 also plays important roles in cellular senescence and p53 regulation (35). Specifically, phospho-PRAS40 decreases p53 expression level (35) and inhibits pro-apoptotic gene expression (36). Our data suggest that quercetin blocks signaling through this pathway and therefore may enhance p53 stability. Additionally, we found that quercetin-treated eSCs show a reduction in ERK1/2 phosphorylation (Figure 6A-D).

ERK1/2 is a parallel signaling pathway to the AKT pathway and downstream of receptor tyrosine kinase activation. Here again, inhibition of ERK phosphorylation would lead to a release of suppression of p53. These relationships are summarized in Figure 10.

**Figure 10.**
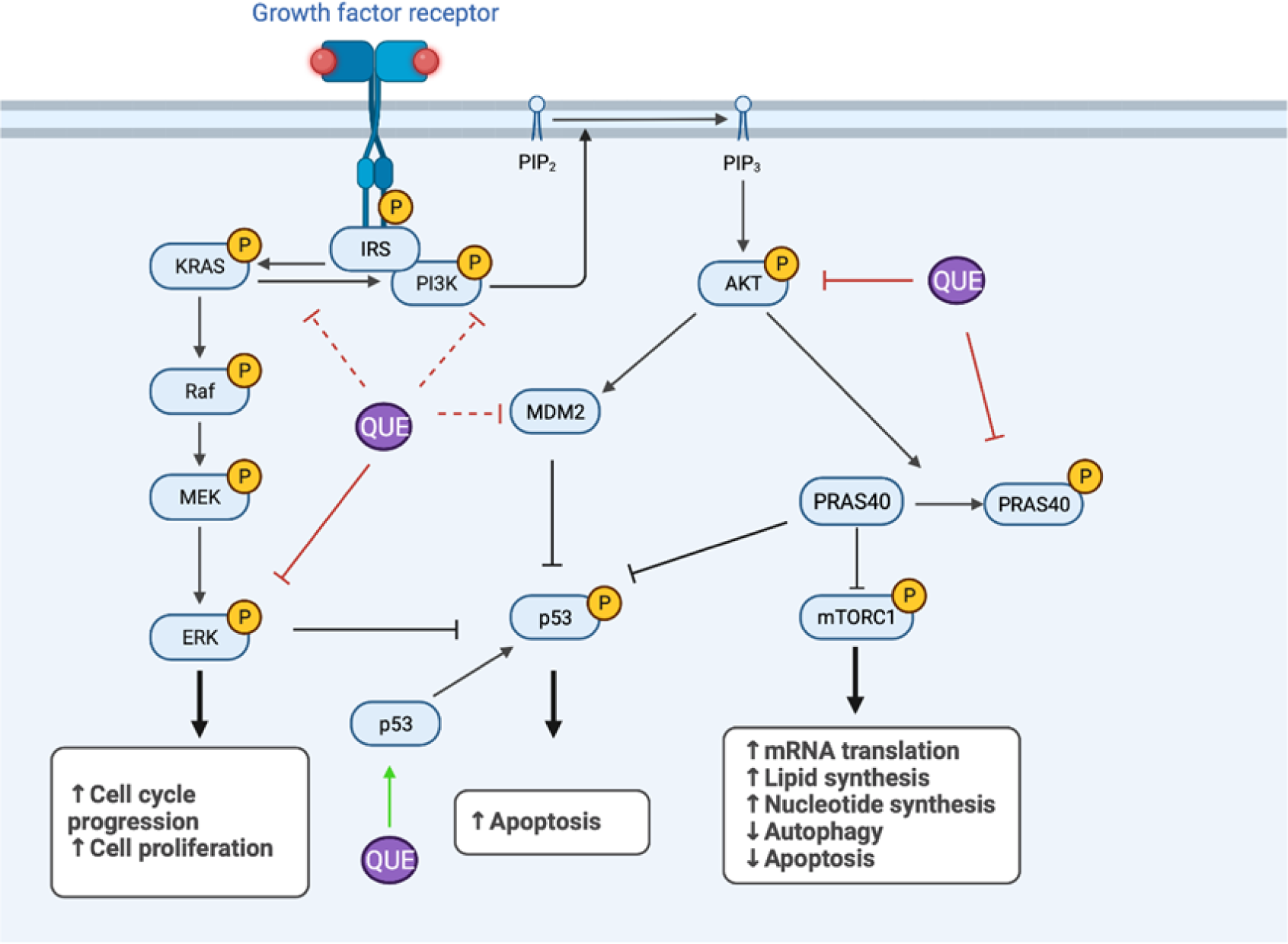
The AKT and ERK1/2 signaling pathways promote cell survival and proliferation and suppress apoptosis. Quercetin (QUE) promotes apoptosis in eSCs by inhibiting the AKT and ERK1/2 pathways and enhancing p53 stability and apoptosis. Green and red solid lines indicate positive and negative effects, respectively, based on our data Dashed green and red lines indicate proposed positive and negative effects, respectively. Created with biorender.com.

Quercetin, a member of a large family of flavonoids found in fruits, vegetables, tea, seeds, nuts, and medicinal botanicals, has been reported to have anti-oxidant, anti-inflammatory, and immunomodulatory activities throughout the body, including the female reproductive tract (37, 38). However, it is the senolytic property of quercetin that has garnered the most attention in recent years. Indeed, recent studies describe the inhibitory effects of eSC senescence and the senescence-associated secretory phenotype (SASP) on decidualization in vitro (39) and this provides a functional link between dysfunctional decidualization, senescence and reduced female fertility (26, 40). Therefore, we searched for evidence that quercetin induces apoptosis of senescent eSCs. As shown in Figure 9, inhibition of apoptosis using Z-VAD, a pan-caspase inhibitor, inhibits decidualization. However, quercetin selectively induces apoptosis eSCs (Figure 9C-D) and specifically eliminates senescent eSCs (Figure 9, panel E). This activity is accompanied by an increase in decidualization and as previously reported, senescent eSCs inhibit decidualization (19). Thus, apoptotic elimination of senescent eSCs may be one mechanism by which quercetin enhances decidualization.

More direct proof of this hypothesis will require isolation and further culture experiments with purified senescent stromal cells. The hallmarks of cellular senescence are cell cycle arrest caused by increased expression of p16 and p21, changes in the nuclear membrane (loss of lamin B1), the production of SASP factors, and the expression of SA-β-gal. However, there are no global characteristics that identify senescent cells in all tissues, and that can be easily applied at the single cell level. This is a major challenge for studies of cellular senescence. Markers of senescence tend to be different in various cell types and there is no standard gene or protein expression pattern that can definitively identify these cells across tissue types. However, increased p16 and p21 expression and reduction of lamin B1 protein are consistent with a senescence phenotype in a subset of our stromal cells as shown in Figure 8D after exposure to H_2_0_2_ (a common method to induce senescence) and the expression of SA-β-gal by larger eSCs shown in Figure 9E-F.

Of course in a larger context, the roles of senescence in aging, cell differentiation, as well as tissue injury and repair have been widely reported (41–43). Senolytic agents have been developed to treat aging-related and other conditions by selectively clearing senescent cells (i.e., growth arrested, viable cells that have undergone metabolic and gene expression changes) (3). In the context of reproduction, quercetin supplementation improves fecundity in young female mice (44) and increases pregnancy rates in a small cohort of patients with polycystic ovary syndrome (PCOS) (45). Additionally, quercetin administration significantly reduces the growth of endometrial implants in rats (4) and mice (5). However, in these studies there is no direct evidence that the effects of quercetin are mediated by its senolytic activity.

Together, these findings warrant future studies to explore the role of senescence in endometriosis and the use of quercetin and other senolytic agents as potential therapies or adjunct treatments for endometriosis and associated infertility. Given the low risk of quercetin as a common dietary constituent, we propose that quercetin treatment should also be tested in clinical trials for disease prevention in patients at risk for endometriosis, where decidualization defects and senescent phenotypes could be monitored through the longitudinal analysis of stromal cells in menstrual effluent.

## Material and methods

### Materials

Quercetin was purchased from Alfa Aesar/Fisher Scientific (Waltham, MA, US). 8-bromoadenosine 3′,5′-cyclic monophosphate sodium salt (cAMP), IBMX (3-isobutyl-1-methylxanthine), forskolin, hydrogen peroxide (H_2_O_2_), medroxyprogesterone acetate (MPA), and *N*-acetylcysteine (NAC) were purchased from Sigma-Aldrich (St. Louis, MO, US). Prostaglandin E2 (PGE2), Z-VAD-fmk, pifithrin-α (PFT), nutlin-3a (Nut), and MK-2206 were purchased from MedChemExpress, LLC (Monmouth Junction, NJ, US).

### Isolation and culture of menstrual effluent (ME)-derived endometrial stromal cells

This study was conducted with approval from the Institutional Review Board (IRB) of Northwell Health (IRB #13-376A). Women of reproductive age (18 to 45 years old) who were not pregnant or breastfeeding or on hormonal contraceptive, and who were menstruating and willing to provide menstrual effluent (ME) samples provided written informed consented. Patients with histologically confirmed endometriosis (documented in a pathology report following laparoscopic surgery and excision of lesions) were recruited and enrolled as ‘endometriosis’ participants (cases) through the ROSE study. Control subjects (controls) who self-reported no history suggestive of a diagnosis of endometriosis were also recruited and enrolled through the ROSE study. No subjects reported having polycystic ovary syndrome (PCOS) or adenomyosis.

Endometrial stromal cells (eSCs) from ME collected from cases and controls were grown, as previously described (15). Briefly, ME-derived eSCs were grown in DMEM containing 10% MSC-fetal bovine serum (FBS), 1% penicillin-streptomycin-glutamine (PSQ) (maintenance media) (Gibco/Thermo-Fisher), and normocin (1:500) (Invivogen, San Diego, CA, US) (maintenance media) at 37°C with 5% CO_2_; cells were split 1:6. Passage 1-3 eSCs were used for experiments (unless otherwise indicated) or were cryopreserved in 10% DMSO/90% FBS for later use. Formal sample size calculations were not performed, as the study of ME-derived stromal cells does not involve ethical, time or cost issues which warrant sample size calculations (46). Sample sizes for functional cell-based assays were consistent with our prior studies (14, 15).

### eSC proliferation assays

ME-derived eSCs were plated in 96-well plates (100 µL/well, 1.5X10^4^/mL) in maintenance media. The next day, media was aspirated and replaced with DMEM containing 2% FBS+1% PSQ and normocin (1:500) (assay media) and treated with vehicle or quercetin (0-50µM) (N=4-6 per condition) at 37°C with 5% CO. After a 72 hr incubation, cells were washed once with cold PBS, aspirated, and frozen at −80°C until assayed for cell proliferation using the CyQUANT Cell Proliferation Assay, according to the manufacturer’s directions (Thermo-Fisher, Waltham, MA, US). CyQuant dye binds to DNA, and the fluorescence emitted by the dye is linearly proportional to the number of cells in the well. Data are shown as relative cell number for each subject’s data point, with each group mean±SD indicated for quercetin concentrations (0-50µM).

### Decidualization assays using ME-derived cultured eSCs

In the morning, confluent monolayers of eSCs (passages 1-3) isolated/grown from controls and endometriosis cases were lifted with trypsin/EDTA, washed, resuspended in maintenance media (DMEM, 10% FBS, PSQ, and normocin), and plated in 96-well plates (100 µL/well, 2.5X10^5^/mL). The same evening, media was aspirated and replaced with decidualization assay media (DMEM, 2% FBS, PSQ, and normocin). After an overnight incubation, cells were treated with either vehicle or quercetin (25μM) for 4 hr (unless described otherwise) before the addition of cAMP (0.5 mM) ± MPA (10^-7^ M) to induce decidualization (N=3-5 per condition). The dose and timing of quercetin addition were based on the literature and optimization studies using eSCs obtained from control subjects. After 48 hr (following cAMP±MPA treatment), supernatants were collected and frozen at −80°C until assayed for insulin growth factor-binding protein 1 (IGFBP1) or prolactin (PRL) proteins by ELISA (R&D Systems, Minneapolis, MN), as previously described (15). Protein data for decidualization assays are shown as IGFBP1 or PRL protein (pg/mL) found in cell-free culture supernatants after 48 hr, unless otherwise specified.

In a subset of samples, decidualization was analyzed by measuring *IGFBP1* and *PRL* mRNA expression by quantitative reverse transcription PCR (qRT-PCR). Briefly, high quality RNA was isolated from cells using the RNeasy Universal Plus Mini Kit (Qiagen). RNA quality was analyzed using the NanoDrop spectrophotometer (Wilmington, DE, US) and the Bioanalyzer (Agilent Technologies Genomics, Santa Clara, CA, US); samples with OD260:280 and OD260:230 ratios >1.9 were converted to double stranded cDNA using the High-Capacity cDNA Reverse Transcription Kit (Applied Biosystems, Foster City, CA, US). Real time qPCR (RT-qPCR) reactions using specific primers and the Roche Universal Probe Library (see Supplementary Table S1) were performed in triplicate using the Eurogentec qPCR MasterMix Plus (AnaSpec, Inc, Fremont, CA, US) and the Roche LightCycler 480. Relative changes in gene expression were calculated as fold-changes using the comparative 2^(-ΔΔCt) method; based on optimization studies, human *HPRT* was used as the housekeeping gene for normalizing transcript levels, as previously described (15).

The effects of MK-2206 (AKT inhibitor), pifithrin-α (p53 inhibitor), nutlin-3a (p53 activator), and Z-VAD (pan-caspase inhibitor) on decidualization were examined as described above, except vehicle, MK-2206 (1μM), Z-VAD (40μM) or PFT (40μM) was added to eSC cultures with vehicle or quercetin (25μM); 4 hr later cells were treated with cAMP alone or cAMP±MPA and processed as described above. Nutlin-3a (100nM) was added 24 hr following cAMP+MPA treatment. Doses were chosen based on the literature and results of in-house optimization studies with eSCs where cytotoxicity (assessed by neutral red(47)) was not detected.

### Immunofluorescent staining for IGFBP1 and assessment by confocal microscopy

Control-eSCs grown to confluence on glass cover slips in 24 well plates in maintenance media were switched to assay media (DMEM, 2% FBS, PSQ). The next day, cells were treated with vehicle or quercetin (25μM) for 4 hr prior to the addition of cAMP+MPA or vehicle, as described above for decidualization assays. After 36 hr, cells were treated with GolgiPlug (BD Biosciences, San Jose, CA) to block IGFBP1 release and then 12 hr later (48 hr after vehicle or cAMP+MPA), cells were washed and fixed with 4% formaldehyde (freshly prepared), permeabilized/blocked with 5% mouse serum/0.3% Triton X-100/1X PBS, and stained with anti-IGFBP1-AF647 antibody (Santa Cruz, sc-55474 AF647, 1:100) and then stained with Alexa488-phalloidin (Abcam, 1:5000) and washed and mounted with Antifade reagent with DAPI (Fisher Scientific). Cells were then examined under a confocal microscope (Zeiss LSM900) using the 20X objective.

### Assessment of intracellular cAMP production by eSCs following forskolin or quercetin

Intracellular cAMP ([cAMP]_i_) production by eSCs isolated from controls and endometriosis cases was analyzed using the cAMP ELISA kit (Cayman Chemical, Ann Arbor, MI, US) according to the manufacturer’s guidelines. Briefly, eSCs were grown to confluence and then seeded into 6-well plates at 3×10^5^ cells/mL (2 mL/well) in maintenance media. 6 hr later, maintenance media was aspirated and replaced with 2% FBS-containing assay media. After an overnight incubation, eSCs were treated with IBMX (0.1mM) for 30 min, followed by either vehicle, forskolin (25µM), or quercetin (25µM, as indicated) and incubated at 37°C with 5% CO_2_ using a method that was optimized in pilot studies. After 20 min, the media was aspirated and the cells were washed once with ice cold PBS and lysed with 0.25 mL 0.1N HCl for measuring [cAMP]_i_, as per the manufacturer’s directions. All samples were diluted 1:2 with ELISA diluent for the cAMP ELISA assay. Data are expressed for each subject’s cells as cAMP pMol/mL with the mean±SD for the group.

### Measurement of basal and H_2_O_2_-induced oxidative stress using eSCs

Control- and endometriosis-eSCs isolated from controls were plated in black 96-well plates with clear bottoms (100 µL/well, 2.5X10^5^/mL) in maintenance media. Media was aspirated and replaced with assay media and after an overnight incubation, cells were washed with HBSS, labeled with DCF-DA (20 µM) in HBSS for 30 min, and washed with HBSS 1% FBS. Phenol red-free DMEM containing 2% FBS + PSQ (100µL/well) was then added, and cells were treated with either vehicle or quercetin (25µM) (or NAC, 10mM) and incubated at 37°C with 5% CO_2_ for 3 hr. Then cells were either treated with vehicle or H_2_O_2_ (500µM) and analyzed for oxidative stress 3 hr later in a quantitative manner using the VICTOR3 fluorescence plate reader (Perkin-Elmer) at an excitation wavelength of 485nm and emission wavelength of 535nm. This assay measures the conversion of non-fluorescent DCFH-DA into fluorescent dichlorofluorescein (DCF) by reactive oxygen species (ROS) intermediates (48). Data are presented for each subject’s cells as percent vehicle-control oxidative stress, with the mean±SD for each group (control vs. endometriosis case) according to condition (basal vs. induced).

### Blocking the effect of H_2_O_2_-inhibition of decidualization

Testing the effect of quercetin on H_2_O_2_-impaired decidualization by eSCs was performed as described above, except eSCs in maintenance media were pre-treated with either vehicle or 250µM H_2_O_2_ for 2 hr and then washed. Maintenance media was replaced with 2% FBS-assay media. 48 hr later, eSCs were treated with vehicle or quercetin (25µM) for 4 hr and then stimulated with cAMP±MPA to induce decidualization and processed as above.

### Proteome profiler array

Confluent eSCs were plated in maintenance media in 100 mm plates and then switched to assay media 6 hr later. The next day, eSCs were treated with vehicle or quercetin (25µM) for 4 hr and then the cell lysates were processed and analyzed, as described by the Human Phospho-Kinase Array Panel Kit (ARY003C, R&D Systems). Image spots were quantified according to the manufacturer’s protocol using ImageJ.

### Western blotting

Control-eSCs or endometriosis-eSCs were plated in maintenance media in 15 mm plates and, when confluent, the media was replaced with 2% FBS-assay media. The next day eSCs were treated with vehicle or quercetin (25 µM) for 4 hr. Cell pellets were resuspended in lysis buffer containing protease and phosphatase inhibitors (Halt™ Protease and Phosphatase Inhibitor Cocktail, Thermo Fisher #PI78442) and incubated on ice for 25 min, followed by centrifugation at 13,000 rpm for 10 min at 4°C. Supernatants were collected and protein levels were quantified using the Pierce BCA Protein Assay Kit (Thermo Fisher, #23225); up to 65 µg/lane was run on 4-12% NuPAGE Bis-Tris 1.5 mm gels in NuPAGE MOPS running buffer (Invitrogen). Proteins were transferred onto Immobilon FL-PVDF membranes, which were blocked at room temperature for 1LJhr in PBS 0.1% Tween-20 (PBST) and 5% nonfat milk powder (#9999 CST). Membranes were rinsed in PBST and incubated overnight at 4°C in primary antibody diluted in PBST containing 5% bovine serum albumin or 5% milk powder, depending on the manufacturer’s recommendation (antibodies are listed in Supplementary Table S2). The next day, membranes were washed in PBST and incubated for 1LJhr at RT in HRP-conjugated secondary antibodies diluted in PBST containing 5% nonfat milk. Membranes were washed again in PBST and imaged by the addition of Clarity ECL substrate (#1705060, BioRad) or SuperSignal West Dura Extended Substrate (#34075, Thermo Fisher) on a ChemiDoc system (BioRad). SeeBlue Plus2 Pre-stained molecular weight standard (#LC5925, Thermo Fisher) was used to estimate protein weight. Western blots were stripped with ReBlot Plus (#2504MI, Millipore), according to the manufacturer’s suggestions, and then re-blocked and probed as described above.

### Assessment of senescence by western blotting

Briefly, eSCs were plated as described for western blotting above, except eSCs (p2-p3) in assay media were treated with vehicle or H_2_O_2_ (250 µM) for 2 hr. After the media was aspirated with replaced with assay media (DMEM 2% FBS, PSQ), eSCs were incubated for 2, 4, 6, or 8 days (as indicated) and then cell lysates were processed as described for western blotting using the Senescence Marker Antibody Sampler Kit (#56062, Cell Signaling Technology). For some experiments, eSCs were treated with vehicle or quercetin (Q, 25μM) for 4 hr post-H_2_O_2_ and analyzed for senescence markers at indicated times.

### scRNA-Seq analysis of fresh menstrual effluent

Single-cell RNA sequencing (scRNA-Seq) data from whole, fresh ME, as previously described (18), was re-analyzed to determine *ADCY1-10* expression by ME-associated eSCs from control subjects.

### Assessment of apoptosis and senescence by flow cytometry

#### Apoptosis

Confluent cultures of control-eSCs (p3-p4) grown in maintenance media were switched to assay media and the following morning they were treated with vehicle or quercetin (25µM) for 24 hr or 48 hr (as indicated) prior to harvesting and processing for analysis of apoptosis by flow cytometry using FITC-Annexin V Apoptosis Detection Kit (BD Biosciences), according to the manufacturer’s directions. Flow cytometry data (fold-change after quercetin treatment) was analyzed on a BD Symphony™ Flow Cytometer and using FlowJo™ v10 software.

#### Senescence

Confluent cultures of control-eSCs (p3-p4) were prepared as described above, except they were processed for fluorometric β-galactosidase staining using DDAO (9H-(1,3-dichloro-9,9-dimethylacridin-2-one), as previously described using the BD Symphony™ Flow Cytometer and FlowJo™ v10 software. The percentage of large senescent cells (as measured by forward scatter, FSC-A) among total viable cells per sample was calculated.

#### Statistics

Analyses and graphical presentations were performed using the GraphPad Prism 5.0 software. The results are presented as the datum for each subject’s cells with the meanLJ±LJstandard deviation (SD) shown for each group. ANOVA with appropriate post-tests for multiple comparisons. Unless indicated, two groups were compared using Wilcoxon matched-pairs signed-rank test – or by Student’s t test for unpaired data (as indicated). P values of 0.05 or less were considered significant (*pLJ<LJ0.05; **pLJ<LJ0.01; ***pLJ<LJ0.001; ****p<0.0001). All data are available by request to the corresponding authors.

## Author contributions

Conceptualization: JD, CNM, PKG, PS-T. Experimentation and data collection: JD, XX, PKC, NH, AJS, RPA, PS-T. Formal analysis: PKG, CNM, JD, AJS. Cell maintenance: XX, PKC. Funding acquisition: CNM, PKG. Writing: original draft: JD, CNM, PKG. Writing: review and editing: JD, CNM, PKG, PKC, RPA, AJS. All authors read and approved the final manuscript.

## Data Availability

All data produced in the present study are contained in the manuscript and are available upon reasonable request to the authors

https://www.ncbi.nlm.nih.gov/geo/query/acc.cgi?acc=GSE203191

## Acknowledgements

The authors would like to thank the coordinators of the ROSE study – Margaret DeFranco, Kristine Elmaliki (now Sadicario), and Amber LaGuerre – for their help in recruiting and enrolling research subjects who donated menstrual effluent for the study. We would like to thank all research subjects who participated in the ROSE study, as well as Laurianna Frasson and Rixsi Herrera for processing ME and isolating/freezing endometrial stromal cells. In addition, we would like to thank Michael Ryan and the team in the Biorepository for maintaining and tracking cryopreserved endometrial stromal cells, as well as Amanda Chan, PhD, who assisted with the microscopy studies. Additionally, we would like to thank JD’s thesis committee members: Drs. Kim Simpfendorfer, Philippe Marambaud, and Myoungsun Son for their research insights and advice over the past two years.

## Funding

This work was supported by the Northwell Health Innovations Award and the Endometriosis Foundation of America

---

**Supplementary Figure S1.**
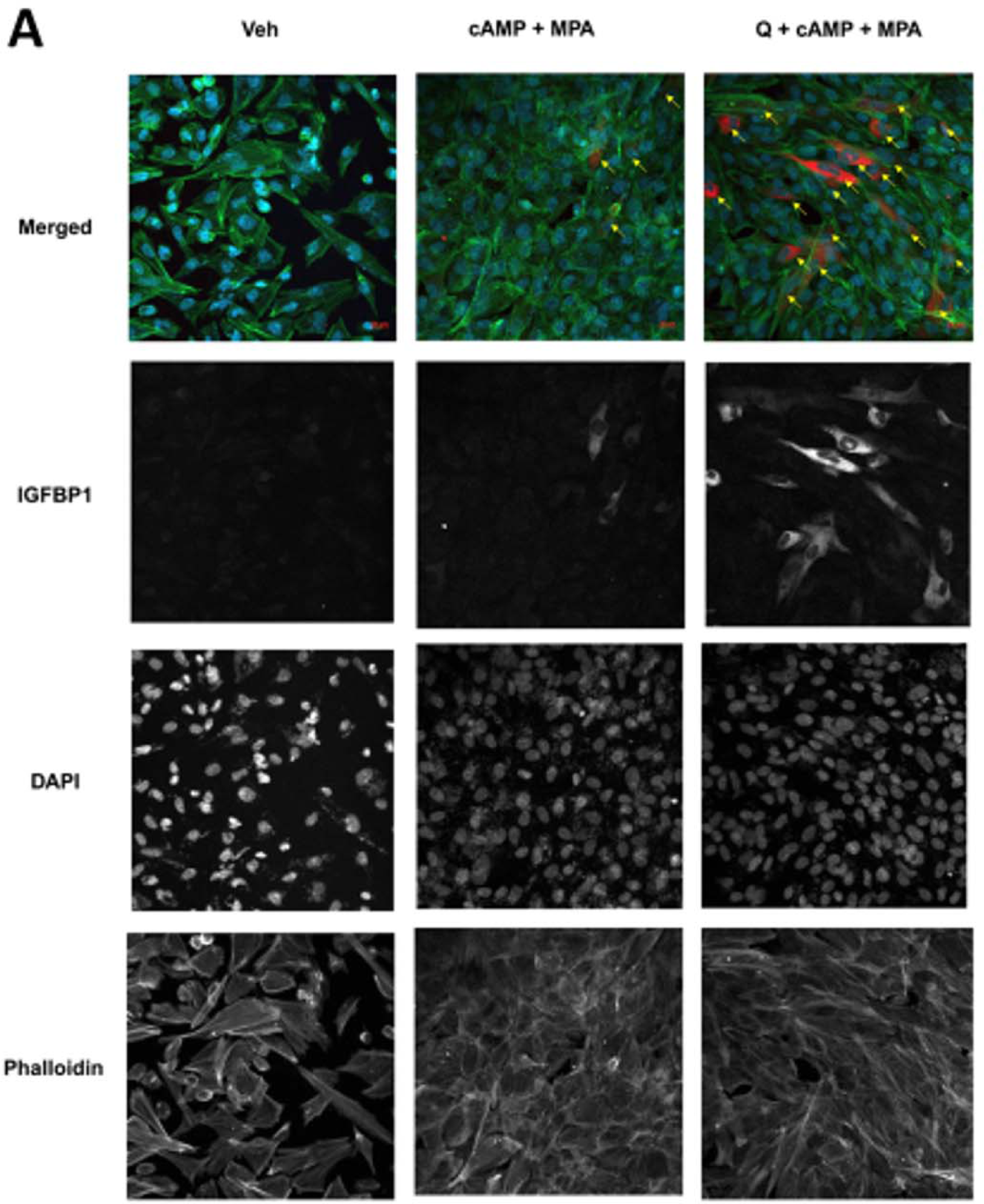
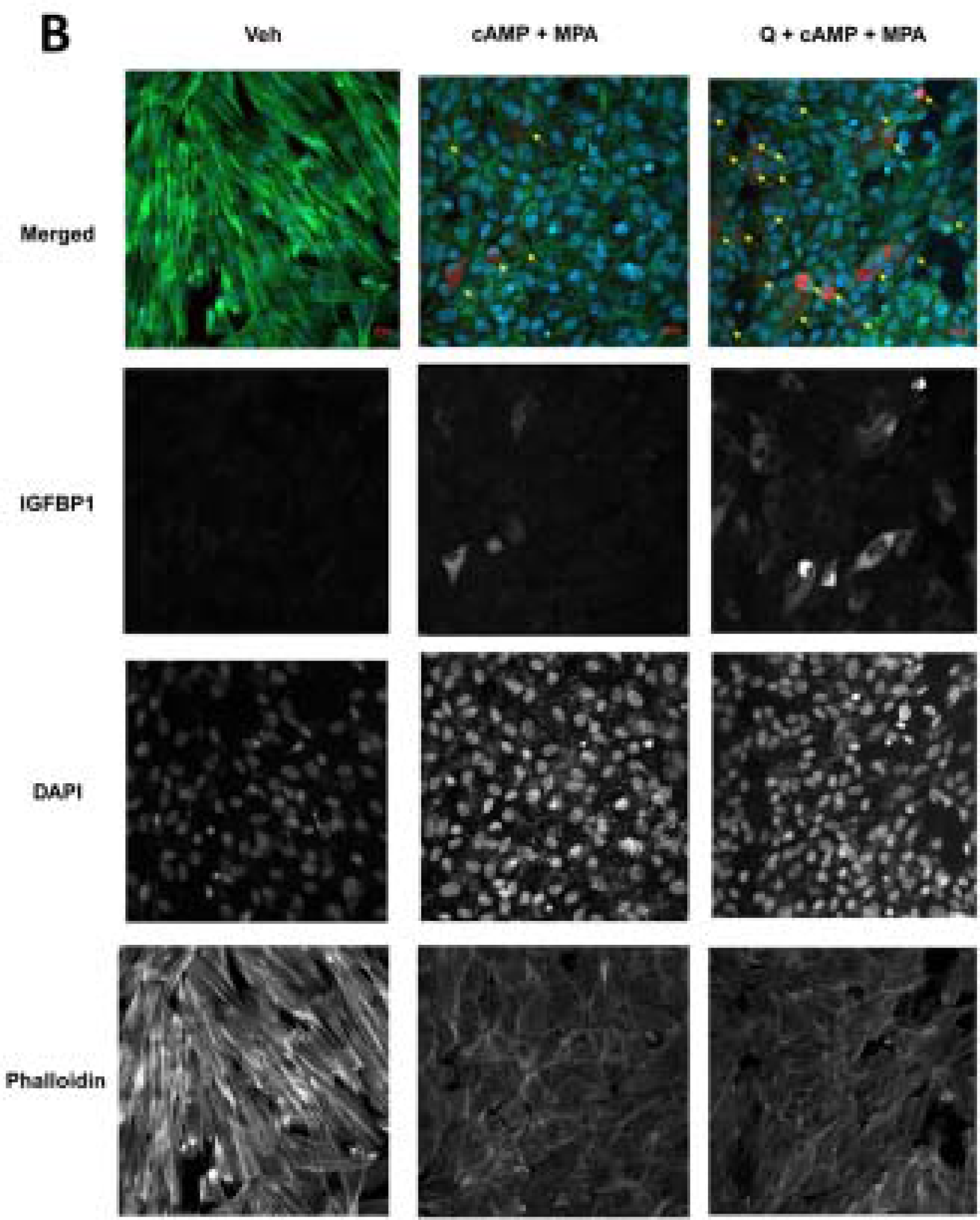
Quercetin enhances decidualization by control ME-eSCs as determined by immunofluorescence. Control-eSCs (Control #1 (A) Control #2 (B)) were treated with vehicle or quercetin for 4 hr followed by vehicle or cAMP+MPA prior to staining and confocal imaging; images show IGFBP1 (red), phalloidin (green), and DAPI (blue) staining (at 20X magnification). The scale bars (20 µm) are indicated. IGFBP1+ cells are indicated by yellow arrows in the merged images; single channel images are shown for IGFBP1, DAPI, and Phalloidin.

**Supplementary Figure S2.**
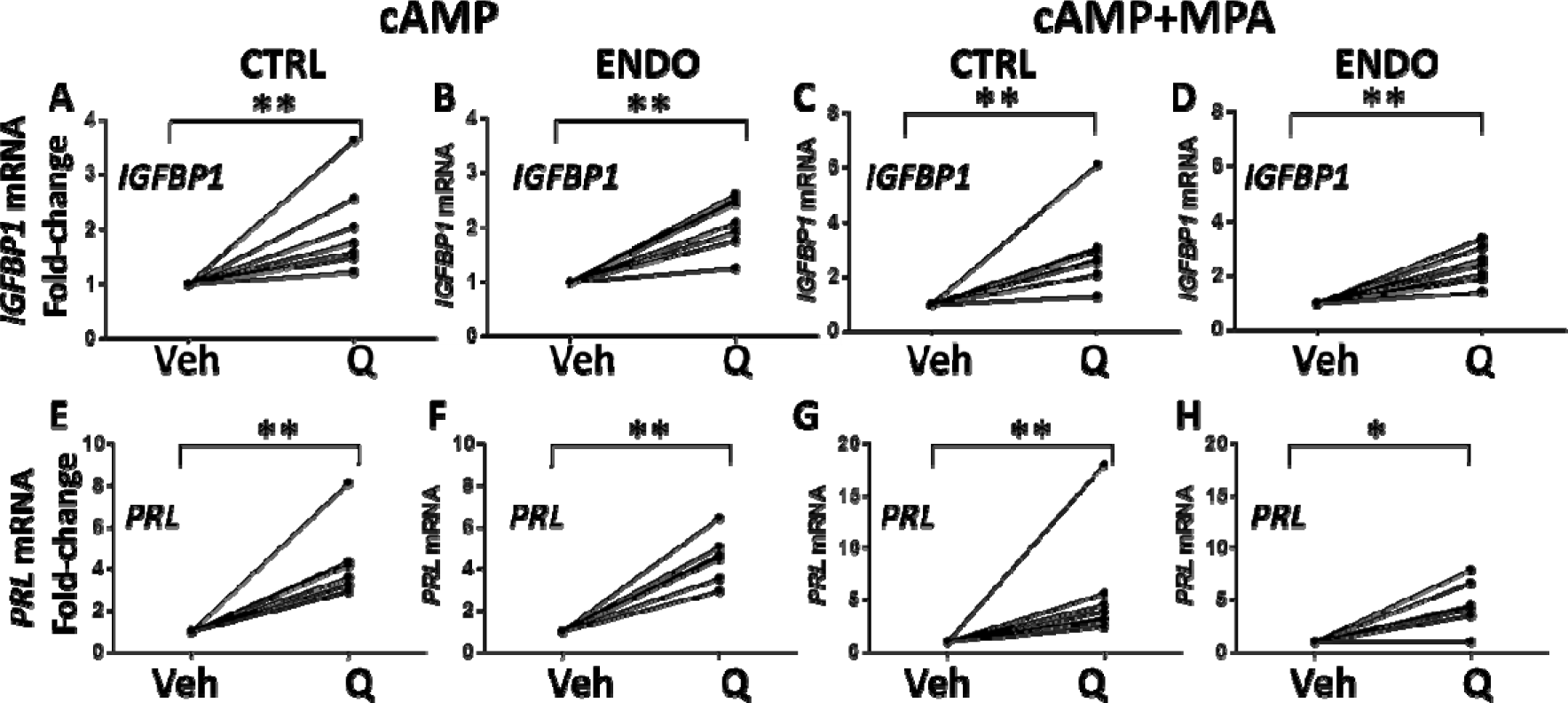
Quercetin enhances decidualization by control-eSCs and endometriosis-eSCs, as determined by *IGFBP1* and *PRL* mRNA production. **(A-H)** Endometrial stromal cells (eSCs) from controls (A, C, E, and G, CTRL) and endometriosis patients (B, D, F, and H, ENDO) were treated with vehicle (Veh) or quercetin (Q, 25_μ_M) for 4 hr prior to the addition of cAMP alone (A-B, E-F) or cAMP+MPA (C-D, G-H). After 48 hr, decidualization was analyzed by measuring *IGFBP1* (A-D) or *PRL* (E-H) mRNA expression by qPCR (shown as fold-change in expression). Data are points connected by a line represent paired mean data points from one individual’s eSCs (±Q). *p<0.05 Veh vs. Q-treated; **p<0.01 Veh vs. Q-treated.

**Supplementary Figure S3.**
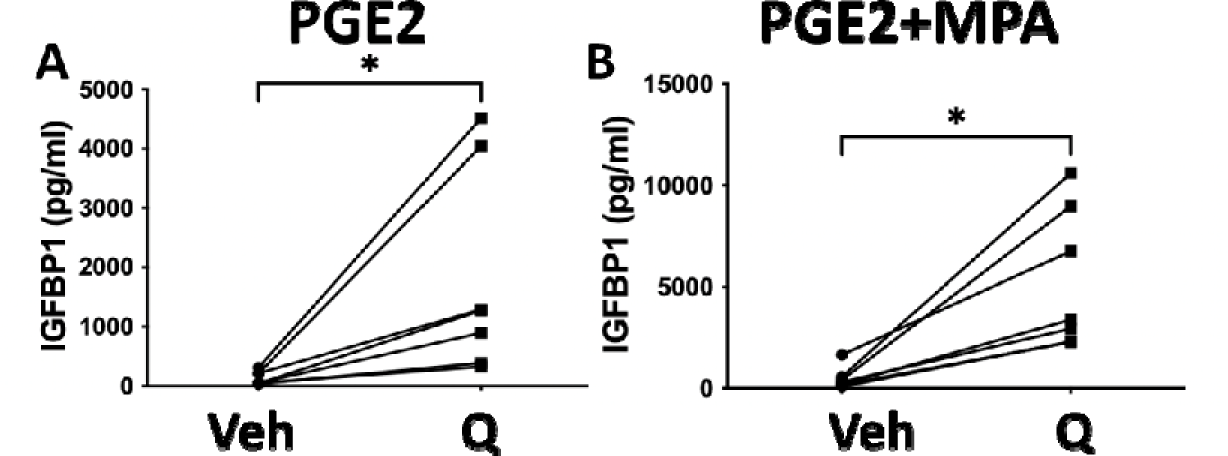
Quercetin enhances PGE2±MPA-stimulated decidualization. **(A B)** Endometrial stromal cells (eSCs) from controls were treated with vehicle (Veh) or quercetin (Q, 25_μ_M) for 4 hr prior to the addition of PGE2 alone (A) or PGE2+MPA (B). Decidualization was analyzed 48 hr later by measuring IGFBP1 protein by ELISA. Data points connected by a line represent paired mean data points from one individual’s eSCs (±Q). *p<0.05 Veh vs. Q-treated.

**Supplementary Figure S4.**
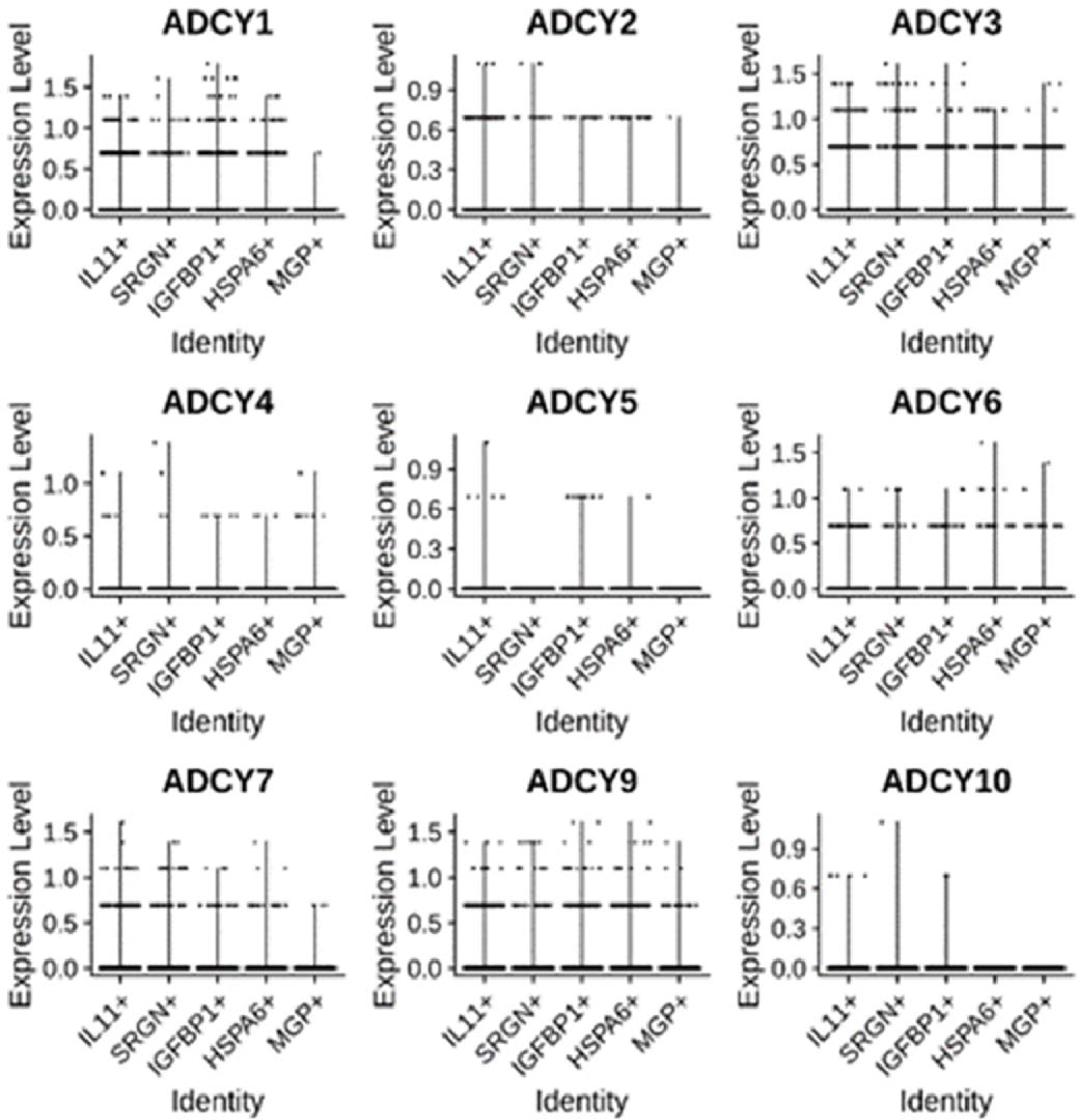
Fresh menstrual effluent (ME) tissue-derived eSCs express multiple *ADCY* transcripts, as shown by scRNA-Seq. *ADCY* gene expression is shown from five different endometrial stromal cell (eSC) clusters (*IL11+, SRGN+, IGFBP1+, HSPA6+,* and *MGP+)* found in menstrual effluent (ME) (18).

**Supplementary Figure S5.**
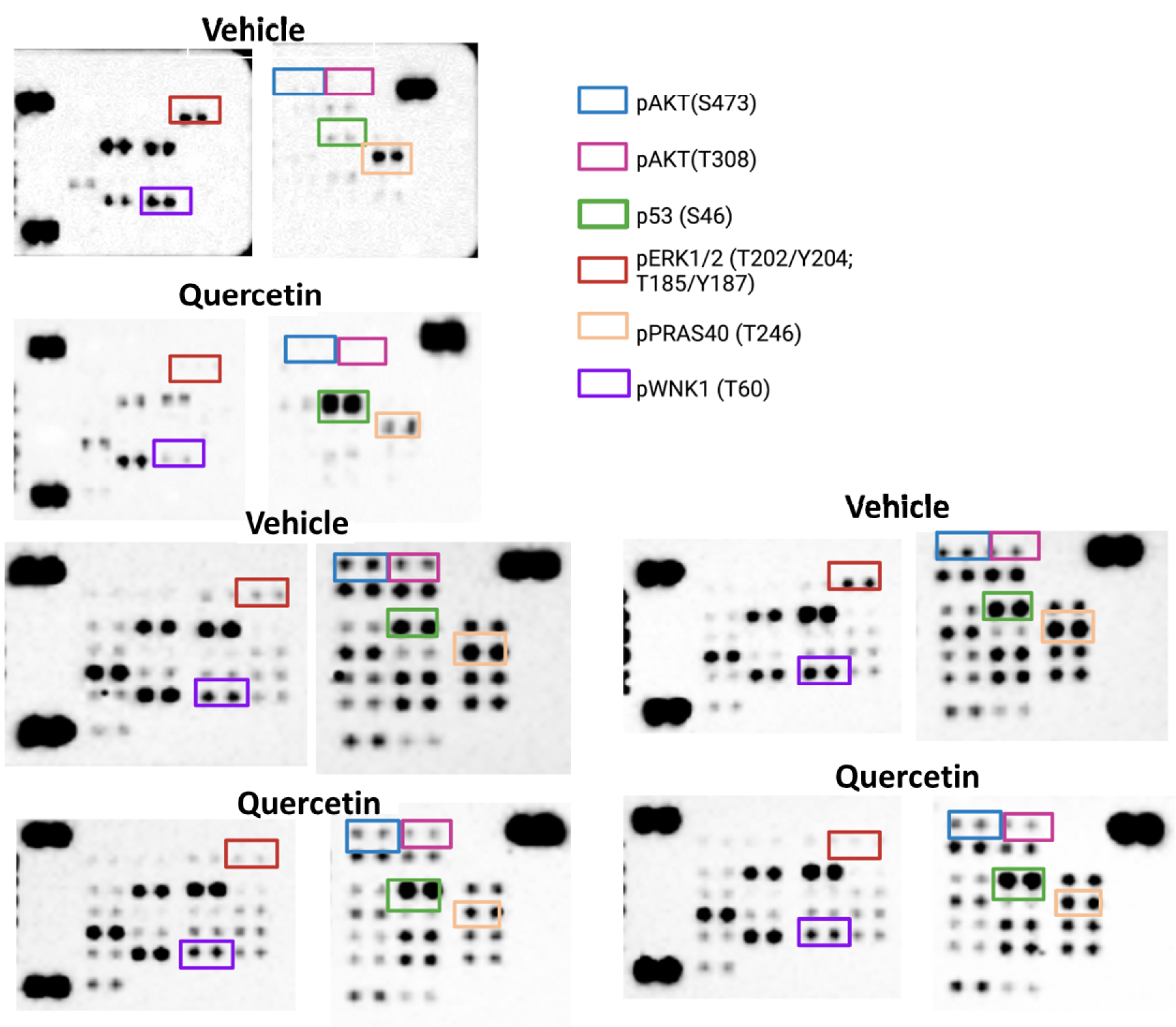
Quercetin inhibits AKT signaling and promotes p53 (Ser46) phosphorylation in eSCs. Control-eSCs were treated with vehicle (Veh) or quercetin (25_μ_M) for 4 hr before analysis using the Phospho-Kinase Array Panel.

**Supplementary Table S1.**
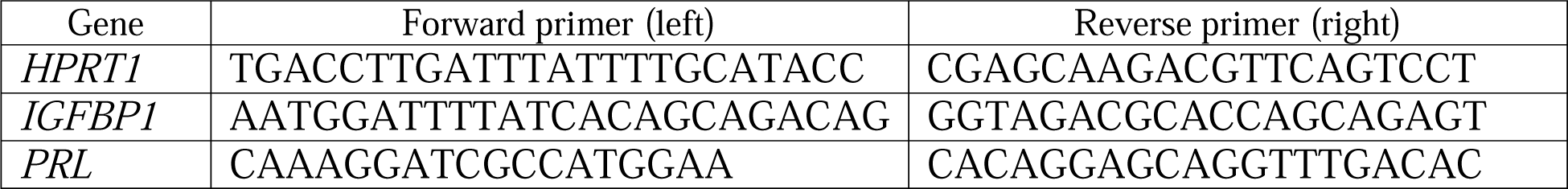
Primers for RT-qPCR.

**Supplementary Table S2.** Antibodies used for western blotting.

| Target | Clone | Dilution | Species raised in | Source |
| --- | --- | --- | --- | --- |
| **Primary antibodies** |  |  |  |  |
| p16 INK4A | Monoclonal | 1:1000 | Rabbit IgG | Cell Signalling Technologie |
| p21 Waf1/Cip | Monoclonal | 1:1000 | Rabbit IgG | Cell Signalling Technologie |
| Lamin B1 | Monoclonal | 1:1000 | Rabbit IgG | Cell Signalling Technologie |
| IL-6 | Monoclonal | 1:1000 | Rabbit IgG | Cell Signalling Technologie |
| MMP3 | Monoclonal | 1:1000 | Rabbit IgG | Cell Signalling Technologie |
| Phospho-AKT (S473) | Monoclonal | 1:1000 | Rabbit IgG | Cell Signalling Technologie |
| AKT | Polyclonal | 1:1000 | Rabbit | Cell Signalling Technologie |
| Phospho-PRAS40 (T246) | Monoclonal | 1:1000 | Rabbit IgG | Cell Signalling Technologie |
| PRAS40 | Monoclonal | 1:1000 | Rabbit IgG | Cell Signalling Technologie |
| Phospho-ERK1/2 (T202/Y204) | Monoclonal | 1:1000 | Rabbit IgG | Cell Signalling Technologie |
| ERK1/2 | Polyclonal | 1:1000 | Rabbit | Cell Signalling Technologie |
| p53 | Polyclonal | 1:1000 | Rabbit | Cell Signalling Technologie |
| GAPDH | Monoclonal | 1:1000 | Rabbit | Cell Signalling Technologie |

## Notes

### Competing Interest Statement

The authors have declared no competing interest.

### Author Declarations

Ethics committee/IRB of The Feinstein Institutes for Medical Research/Northwell Health gave ethical approval for this work (IRB# #13-376A).

### Summary of Updates

We replaced the title to better and more clearly describe the work. We updated the abstract, introduction, and discussion to focus on 'mechanism' and to provide better clarity.

